# DNA methylation changes in glial cells of the normal-appearing white matter in Multiple Sclerosis patients

**DOI:** 10.1101/2021.06.21.21258936

**Authors:** Lara Kular, Ewoud Ewing, Maria Needhamsen, Majid Pahlevan Kakhki, Ruxandra Covacu, David Gomez-Cabrero, Lou Brundin, Maja Jagodic

## Abstract

**Background:** Multiple Sclerosis (MS), the leading cause of non-traumatic neurological disability in young adults, is a chronic inflammatory and neurodegenerative disease of the central nervous system (CNS). Due to the poor accessibility to the target organ, CNS-confined processes underpinning the later progressive form of MS remain elusive thereby limiting treatment options. We aim to examine DNA methylation, a stable epigenetic mark of genome activity, in glial cells to capture relevant molecular changes underlying MS neuropathology.

**Methods:** We profiled DNA methylation in nuclei of glial cells, isolated from 38 post-mortem normal-appearing white matter (NAWM) specimens of MS patients (n=8) in comparison to white matter of control individuals (n=14), using Infinium MethylationEPIC BeadChip.

**Findings:** We identified 1,226 significant (genome-wide adjusted *P*-value < 0.05) differentially methylated positions (DMPs) between MS patients and controls. Functional annotation of the altered DMP-genes uncovered alterations of processes related to cellular motility, cytoskeleton dynamics, metabolic processes, synaptic support, neuroinflammation and signaling, such as Wnt and TGF-β pathways. A fraction of the affected genes displayed transcriptional differences in the brain of MS patients, as reported by publically available transcriptomic data. Cell type-restricted annotation of DMP-genes attributed alteration of cytoskeleton rearrangement and extracellular matrix remodelling to all glial cell types, while some processes, including ion transport, Wnt/TGF-β signaling and immune processes were more specifically linked to oligodendrocytes, astrocytes and microglial cells, respectively.

**Conclusion:** Our findings strongly suggest that NAWM glial cells are highly altered, even in the absence of lesional insult, collectively exhibiting a multicellular reaction in response to diffuse inflammation.

## Introduction

Multiple sclerosis (MS) is a chronic inflammatory demyelinating and neurodegenerative disease of the central nervous system (CNS). MS pathology presents with the occurrence of immune-induced demyelinating lesions arising throughout the CNS and translating into various neurological symptoms. The nature of injuries in MS is highly heterogeneous as lesions observe spatio-temporal diversity, i.e. varying across the CNS and disease stages^1^. Disability closely mirrors neuro-axonal deterioration, irrespective of the type of lesions and disease course^2^. While inflammation is unambiguously observed at all stages of the disease, compartmentalized chronic inflammation orchestrated by resident CNS cells dominate later stages of the disease, independently of immune infiltrates or pre-existing demyelination and accounts for the clinical trajectory^3–5^. Ultimately, persistent inflammation and recurrent damage to the myelin insulating axons will eventually exhaust the repair capacity of the nervous tissue^6,7^, a weakened functional recovery presaging transition to the progressive stage of the disease. Whereas major progress has been achieved in understanding and treating early phase of disease development, *via* targeting of peripheral immune cells, the CNS-confined mechanisms underlying the later stage of disease progression remain elusive. This is likely due to the difficulty to gather molecular evidence from the affected tissue itself, brain specimens being accessible post-mortem thereby restricting methodological applications in relatively small case-control cohorts of high-quality samples. This knowledge gap considerably cripples the care of progressive MS forms, leaving clinicians with a scarcity of therapeutic solutions and patients with relentless and untreatable disabilites.

The study of glial cell populations has gained particular interest due to the possibility to decipher the neurotoxic and pro-inflammatory processes precipitating neuronal vulnerability in progressive MS and thereby, to ultimately restore the brain repair capacities favoring remyelination and neuroprotection^8^. Due to their myelin-producing ability, oligodendrocytes provide a structural and trophic support to neurons by controlling the myelination of axons and are particularly abundant in the white matter (WM) compared to the grey matter (GM)^9^. Astrocytes are key homeostatic regulators of the CNS and, as such, exert versatile functions in synapse refinement, neurotransmission, formation of the blood-brain-barrier and metabolic control of the microenvironment, among others, depending on their primary location. Microglia, populating less than 10% of glial cells, are highly responsive CNS-resident macrophages that assume various functions pertaining to their immunocompetent and phagocytic capacities, including but not limited to synaptic pruning, immunosurveillance and clearance. Due to their inherent migratory abilities, microglial cells are highly dynamic and vigilant cells at resting state, continuously surveying their microenvironment^10^. Overall, the CNS homeostasis entails tightly controlled region-specific (e.g. WM *vs*. GM) regulations of cellular phenotypes through multidirectional glia-glia and neuro-glia signaling.

Investigation of the rodent brain in MS-like models and histopathological characterization of post-mortem brain sections of patients have been instrumental in unveiling the overlapping sequences of events occurring in the MS CNS, mostly under lesional immune insult. Emerging evidence from transcriptome studies further support CNS damage in MS to likely ensue from a complex interplay between various glial cell populations, with each cell type manifesting marked spatial and temporal phenotypic heterogeneity^11–15^. Importantly, diffuse abnormalities in myelination and neuroinflammation accumulate outside of the affected areas as well, namely in the Normal Appearing White Matter (NAWM)^16–19^. These changes reflect phenotypic dysfunction of glial cells in the absence of infiltrating leukocytes and macroscopic lesion and have been proposed as prominent early processes preceding newly forming lesions^20–27^. Importantly, alterations of the NAWM have been associated with cortical axonal loss, cognitive decline and clinical disability^28,29^. Yet, the molecular changes occurring in the NAWM remain elusive, most studies capturing highly dynamic transcriptional states of cells from MS lesions. In that regard, exploring the molecular layer of epigenetic changes, which orchestrate both durable and transitional states, could provide additional insight into the mechanisms underpinning brain pathology in progressive MS^30^.

DNA methylation, the most studied epigenetic mark, relies on the stable deposition of a methyl group onto cytosine, primarily in a CpG context, and exerts regulatory action on gene expression that depends on its gene location^31^. Profiling DNA methylation genome-wide at single-base resolution enables probing the chromatin state and genome activity reliably in post-mortem tissue. Comparison of demyelinated and myelinated hippocampi of MS patients has identified aberrant epigenetic changes associated with deregulation a small set of genes^32^. Previous case-control methylome analysis of bulk brain tissue has revealed subtle changes in the white matter of MS patients compared to controls^33^. More recently, by conducting DNA methylation analysis of neuronal nuclei, we have identified functionally relevant changes, including reduced CREB transcription factors activity, associated with neuro-axonal impairment in MS patients compared to controls^34^. Here we aimed to elucidate the molecular alterations occurring in glial nuclei sorted from the NAWM of MS patients in comparison to WM of non-neurological disease control individuals.

## Materials and methods

### Subjects, cohorts and ethics

Brain tissues used for this study and obtained from the Multiple Sclerosis and Parkinson’s Tissue Bank (Imperial College London) were approved by local ethical guidelines. All research included in this manuscript conforms with the Declaration of Heksinki. The material comprises 38 snap-frozen brain tissue blocks collected within 33h post-mortem from NAWM tissue of MS patients (n=8) and WM tissue of controls (n=14) (Table 1). Further details are given in Supplementary Table 1. Control subjects were selected based on a non-neurological cause of death. The samples were further annotated according to brain location following antero-posterior axis and characteristics of the tissue using the human brain atlas sectional anatomy database (http://www.thehumanbrain.info/) prior to dissection. Of note, different brain samples, coming from distinct regions, were used from the same individuals. Samples reaching the following inclusion criteria have been included in DNA methylation analyses: (i) all available samples with sufficient DNA amount from WM-glial nuclei, (ii) samples that passed DNA methylation quality control and (iii) cases with confirmed MS diagnosis and non-neurological controls without any signs of inflammation in the CNS.

**Table 1.**
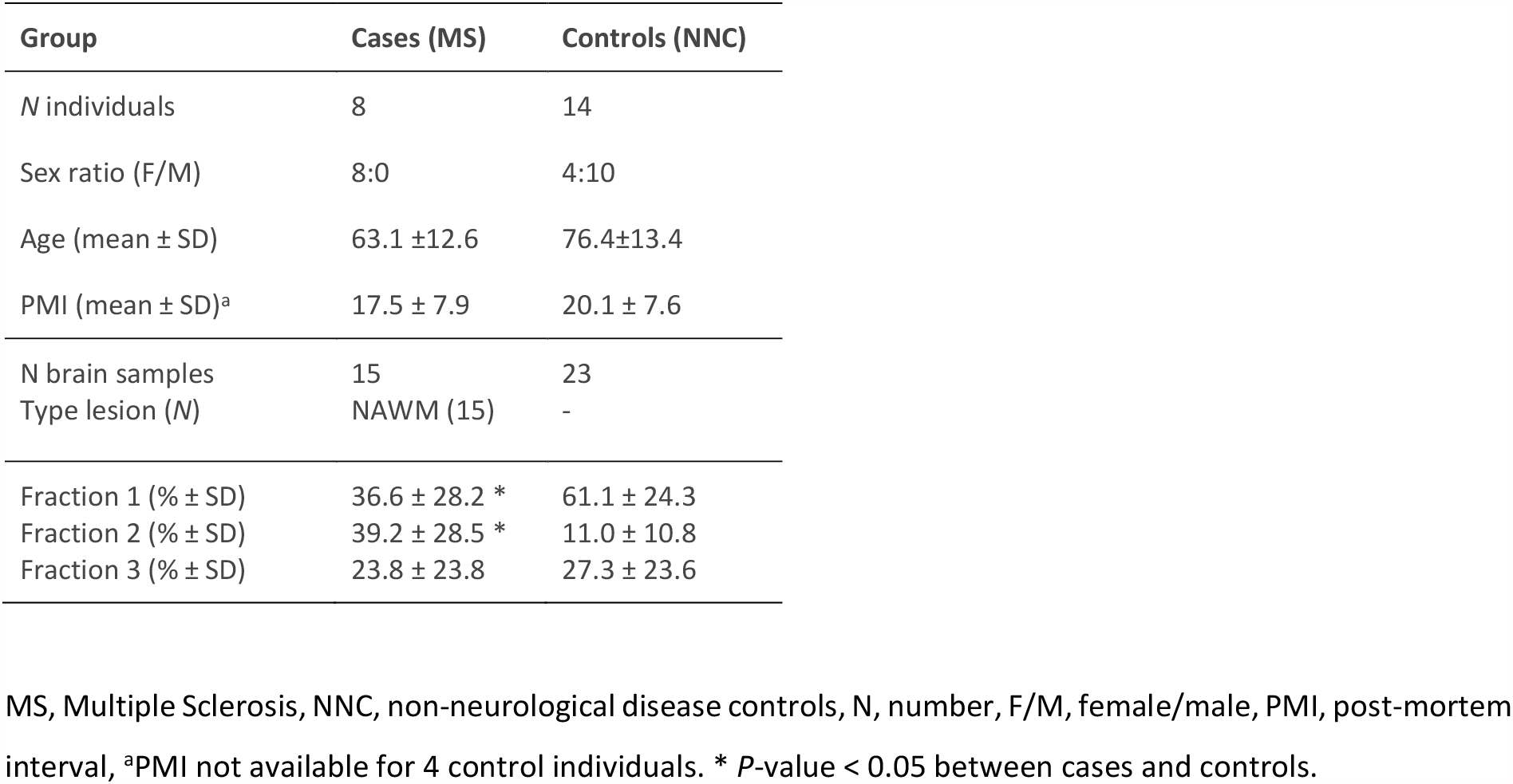
Description of the cohort

### Sample preparation

Fluorescence activated cell sorting (FACS)-based neuronal nuclei isolation from dissected brain tissue was performed as previously described^34^. Briefly, following resuspension of the homogenized brain tissue in hypotonic lysis buffer (0.32M sucrose, 5mM CaCl_2_, 3mM MgAc_2_, 0.1 mM EDTA, 10mM Tris pH.8, 1mM DTT, 0.1 % Triton), nuclei were extracted by ultracentrifugation in sucrose gradient (1.8M sucrose, 3mM MgAc_2_, 1mM DTT, 10mM Tris pH.8) for 2.5 hours at 4°C. Nuclei were further labelled with Alexa Fluor 488 (Invitrogen #A11029)-conjugated anti-NeuN antibodies (1:700, Millipore #MAB377) and separated into neuronal and non-neuronal nuclei by flow cytometry (MoFlo™ high-speed cell sorter). Non-neuronal nuclei were pelleted and stored at −80°C until DNA isolation. Genomic DNA was isolated using QIAmp DNA micro kit (QIAGEN), resuspended in water and stored at −80°C until further use.

### Illumina Human Methylation EPIC

We used Illumina Infinium Human MethylationEPIC BeadChip (Illumina, Inc., San Diego, CA, U.S.A, EPIC) for quantitative and genome-wide DNA methylation profiling. Genomic DNA samples were processed at GenomeScan (GenomeScan B.V., Leiden, The Netherlands), according to manufacturer’s instructions and the BeadChip images were scanned on the iScan system. Samples were randomized ensuring that disease group, gender and age were balanced to control for potential confounding effects. Technicians performing EPIC arrays were blinded to the MS disease status during the experiments.

### DNA methylation analysis

#### Quality control

EPIC data was quality assessed using QC report from the minfi package. All samples passed quality control and were subsequently processed using the Chip Analysis Methylation Pipeline (ChAMP) version 2.9.10^35^ and minfi version 1.24.0^36^ R-packages.

#### Probe filter

Upon loading raw IDAT files into ChAMP, probes were filtered by detection P-value > 0.01, bead count < 3 in at least 5% of the samples, SNPs (minor allele frequency > 1% in European population) and cross-reactivity as identified by Nordlund *et al*.^37^ and Chen *et al*.^38^. After filtering for probes located on X and Y chromosomes, 700,482 probes remained.

#### Between and within-array normalization

Probes were subject to within-sample normalization (Noob), which corrects for two different probe designs (type I and type II probes) included on the EPIC BeadChip. Slide effects, as identified using Principal Component Analysis (PCA), were corrected using empirical Bayes methods^39^ implemented in the ComBat function of the SVA Bioconductor package version 3.26.0.

#### Deconvolution

Reference-free cell type deconvolution was performed using RefFreeEWAS^40^, as no accurate cell based reference models exist for deconvolution of DNA methylation in the glial fraction. The optimal number of fractions for deconvolution was determined by calculating the deviance-boots (epsilon value) over 100 iterations for the range of 1 to 6 fractions, with minimum deviance with 3 fractions. The fractions were obtained by solving the model *Y* = *M* × *Ω* − *T* (where Y = original beta methylation matrix, M = cell type specific beta methylation matrix, Ω = cell proportion matrix and T is number of cell types to deconvolute) using the nonnegative matrix factorization method.

#### Differentially methylated positions (DMPs) and regions (DMRs)

The Limma Bioconductor package version 3.34.9^41^ was used for detection of DMPs with M-values 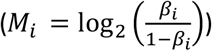 as input as previously recommended^42^. Since several samples with different brain location were used from the same donor, Limma was conducted using within-individual comparions in a linear mixed model (LMM) by estimating the average correlation within individual prior to using the function duplicateCorrelation which was then used in the lmFit step.

The following covariates, as confirmed by PCA, were included in the model: sex, age and cell type proportions. DMRcate version 1.6.53^43^, which identifies DMRs based on kernel smoothing, was applied with default settings (λ=1000, C=2).

#### Gene annotation

Classical EPIC annotations (TSS200, TSS1500, 1stExon, 5’UTR, Gene body, 3’UTR, CGI, Shelf, Shore and Open Sea) as well as Fantom-annotated enhancers were derived from the “IlluminaHumanMethylationEPICanno.ilm10b2.hg19” package. CpG islands (CGIs) were defined as GC content > 50%, observed/expected CpG ratio > 60%, > 200bp while CGI shores and shelves represent 2kb flanking regions within or outside CGIs, respectively. Fisher’s exact test integrated in R version 3.4.3 was used to estimate enrichment (alternative=“greater”) or depletion (alternative=“less”) of features of interest.

### Gene ontology analyses

Gene ontology (GO) analysis of DMPs (P_adj_ < 0.05) was performed using overrepresentation analysis (ORA) from the online software tool WebGestalt (www.webgestalt.org)^44^ under default settings. Findings from ORA were clustered using REVIGO (http://revigo.irb.hr/)^45^ or GeneSetCluster package^46^. STRING network analysis applied to DMPs (P_adj_ < 0.05) was generated using STRING database version 11.0.

## Results

### DNA methylation changes in glial cells from MS patients

We performed genome-wide DNA methylation profiling on bisulfite (BS)-treated genomic DNA isolated from NeuN-negative (hereafter referred to as glial) fraction sorted from the WM of fresh-frozen post-mortem tissue blocks. A total of 38 glial nuclei samples isolated from NAWM tissue of 8 MS patients and WM of 14 non-neurological disease controls (NNC) were profiled using Illumina HumanMethylationEPIC BeadChip (Table 1, Supplementary Table 1). We conducted reference-free deconvolution to account for varying proportions of cell types, which overcomes the current lack of existing reference methylome for distinct human glial cell types, using RefFreeEWAS tool^40^. This method identified optimal separation to be based on three fractions, of which two differ (*P* < 0.05) between MS and NNC samples (Table 1, Supplementary Fig. 1).

After correction for confounders, we identified 1,226 differentially methylation positions (DMPs) mapping to 687 annotated genes, between MS and NNC (adjusted *P*-value, *P*_adj_ < 0.05) (Fig. 1a). Most DMPs (65%, 803/1226) exhibited differences exceeding |Δβ| > 0.05; the most significant ones are listed in Table 2 (complete data is presented in Supplementary Table 2). Slightly more than half of the identified changes (55%, 682/1226) exhibited hypermethylation in MS patients compared to controls. Markedly, a substantial fraction (44%) of the identified DMPs (541/1226) are unique to the Illumina EPIC array, hence not present in the previous Illumina 450K array. These DMPs represented ∼45% (327/687) of the annotated genes, with minor overlap (10/327) with 450K-annotated DMP-genes. Of note, only three and seven DMPs were previously identified in MS vs. NNC bulk brain^33^ and neurons^34^, respectively (Supplementary Figure 2). We examined whether changes clustered in differentially methylated regions (DMRs) and identified 276 DMRs mapping to 225 gene-annotated loci between MS patients and controls (mean Δβ_DMR_ ranging from −0.18 to 0.19). The majority of the DMRs (71%, 197/276) encompassed at least one DMP (Supplementary Table 3).

**Figure 1.**
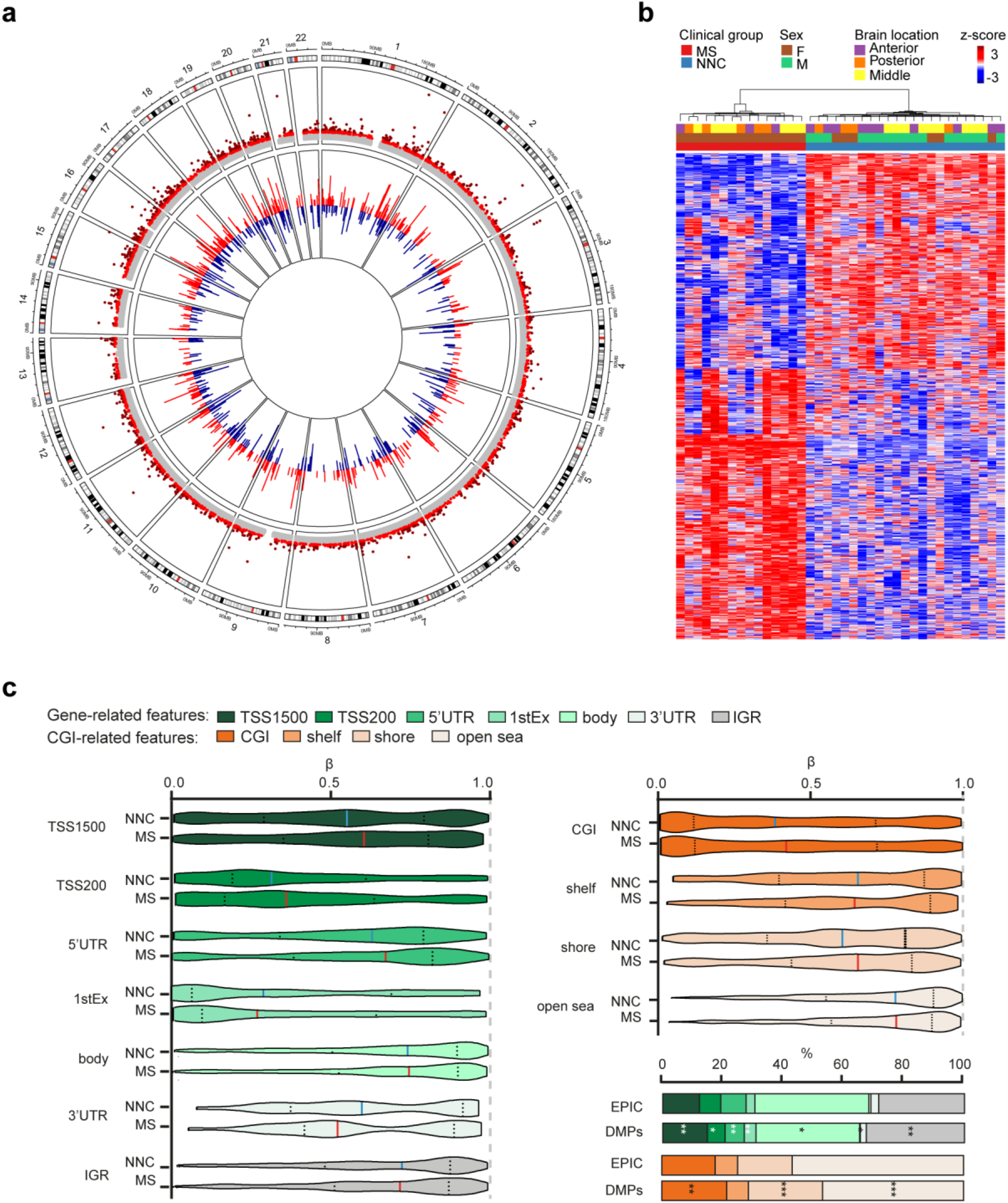
DNA methylation changes in glial nuclei of Multiple Sclerosis patients **a**. Circos plot illustrating differentially methylated CpGs in glial nuclei of Multiple Sclerosis (MS) compared to non-neurological controls (NNC). The outer track is a hg19 ideogram illustrating chromosome and cytoband information. The upper track represents the corresponding −log_10_ (*P*-values), with dark red indicating sites passing significance threshold of Benjamini-Hochberg-adjusted P-value, *P*_adj_ < 0.05 and light red illustrating CpGs with *P* < 0.001. The inner track corresponds to the effect size of DNA methylation changes (Δβ-value) between MS and NNC, with red and blue depicting hyper- and hypomethylation, respectively. **b** Heatmap of the DMPs (*P*_adj_ < 0.05) with red and blue depicting z-score transformed hyper- and hypomethylation, respectively. **c**. Distribution of DMPs (*P*_adj_< 0.05) across classical gene features including promoter-like features (TSS1500, TSS200, 1stExon, 5′UTR), gene body and 3′UTR (shades of green) and CpG island (CGI)-related features including CpGs islands, shores, shelves and open sea (shades of orange). Violin plots depict β-values and barplots DMPs distribution (in comparison to EPIC background). \**P* < 0.05, \*\**P* < 0.001 using Fisher’s exact test for enrichment and depletion analyses.

**Table 2.** Top significant differentially methylated positions associated with Multiple Sclerosis in glial nuclei

| Probe | Chr. | Position | Gene | Feature | cgi | Mean MS | Mean NNC | $\Delta\beta$ | *P* | *P*.adj |
| --- | --- | --- | --- | --- | --- | --- | --- | --- | --- | --- |
| cg20810195 | 1 | 86970291 |  | IGR | opensea | 0.738 | 0.868 | -0.114 | 1.7E-11 | 4.6E-07 |
| **cg09674340** | 1 | 202509286 | PPP1R12B | 1stExon | opensea | 0.933 | 0.845 | 0.091 | 1.5E-15 | 6.4E-11 |
| cg26919182 | 1 | 202522232 | PPP1R12B | Body | opensea | 0.935 | 0.598 | 0.337 | 3.7E-33 | 2.6E-27 |
| **cg15817705** | 1 | 209406063 |  | IGR | shore | 0.737 | 0.849 | -0.104 | 3.1E-11 | 8.2E-07 |
| cg12691488 | 1 | 243053673 |  | IGR | island | 0.134 | 0.297 | -0.152 | 2.0E-12 | 6.6E-08 |
| cg15228509 | 1 | 243074717 |  | IGR | opensea | 0.840 | 0.555 | 0.292 | 2.4E-28 | 8.3E-23 |
| **cg19765154** | 2 | 191524409 | NAB1 | Body | opensea | 0.715 | 0.885 | -0.177 | 1.0E-25 | 2.4E-20 |
| **cg20262915** | 2 | 191524489 | NAB1 | Body | opensea | 0.395 | 0.489 | -0.104 | 4.7E-12 | 1.4E-07 |
| cg06759085 | 2 | 191555076 | NAB1 | 3'UTR | opensea | 0.780 | 0.895 | -0.109 | 6.3E-16 | 3.1E-11 |
| cg11532947 | 2 | 191555480 | NAB1 | 3'UTR | opensea | 0.890 | 0.939 | -0.047 | 1.4E-12 | 5.0E-08 |
| cg00148935 | 3 | 16398839 | RFTN1 | Body | opensea | 0.881 | 0.646 | 0.233 | 1.1E-20 | 9.2E-16 |
| cg11643285 | 3 | 16411667 | RFTN1 | Body | opensea | 0.954 | 0.837 | 0.117 | 1.7E-23 | 3.0E-18 |
| cg17238319 | 3 | 16428391 | RFTN1 | Body | opensea | 0.937 | 0.753 | 0.184 | 2.6E-22 | 3.1E-17 |
| **cg08034535** | 4 | 140375469 | RAB33B | 1stExon | island | 0.109 | 0.057 | 0.049 | 6.3E-12 | 1.8E-07 |
| **cg26516287** | 7 | 12629275 | SCIN | Body | opensea | 0.875 | 0.810 | 0.064 | 2.9E-11 | 7.9E-07 |
| **cg06513015** | 7 | 64459246 | ERV3-1 | 5'UTR | opensea | 0.864 | 0.681 | 0.190 | 8.2E-20 | 6.4E-15 |
| **cg07852945** | 9 | 84303915 | TLE1 | TSS1500 | island | 0.230 | 0.080 | 0.144 | 8.4E-18 | 4.9E-13 |
| cg02716779 | 9 | 131016143 | DNM1 | 3'UTR | shelf | 0.785 | 0.679 | 0.108 | 8.0E-16 | 3.7E-11 |
| **cg25294185** | 11 | 65487814 | RNASEH2C | Body | island | 0.022 | 0.136 | -0.116 | 1.6E-21 | 1.6E-16 |
| cg09241427 | 11 | 70860363 | SHANK2 | 5'UTR | opensea | 0.906 | 0.817 | 0.095 | 1.4E-12 | 5.1E-08 |
| **cg09516963** | 12 | 68042445 | DYRK2 | TSS200 | island | 0.676 | 0.317 | 0.377 | 1.2E-18 | 7.4E-14 |
| **cg25438338** | 14 | 24701525 | GMPR2 | TSS1500 | shore | 0.166 | 0.091 | 0.075 | 2.1E-12 | 6.6E-08 |
| **cg23719534** | 15 | 101099284 |  | IGR | island | 0.969 | 0.880 | 0.093 | 6.6E-19 | 4.6E-14 |
| **cg04946709** | 16 | 59789030 | LOC644649 | Body | island | 0.659 | 0.814 | -0.149 | 2.2E-15 | 9.0E-11 |
| **cg20299935** | 17 | 21795943 |  | IGR | opensea | 0.812 | 0.692 | 0.122 | 4.5E-13 | 1.7E-08 |
| **cg08906898** | 20 | 34319899 | RBM39 | Body | opensea | 0.767 | 0.906 | -0.130 | 3.9E-17 | 2.1E-12 |
| **cg17612569** | 21 | 27107221 | GABPA | TSS200 | island | 0.047 | 0.219 | -0.170 | 7.3E-23 | 1.0E-17 |

Ch, chromosome, cgi, CpG island, NNC, non-neurological control, MS, Multiple Sclerosis, $\Delta\beta$ , delta-beta values.
Probes highlighted in bold are included in differentially methylated region.

We next explored the distribution of DNA methylation changes (DMPs, *P*_adj_ < 0.05) according to gene- and CpG island (CGI)-related features and found overall higher β-values at DMPs in MS compared to controls across promoter-related features (TSS1500, TSS200, 5’UTR) while lower methylation was observed in the first exon and 3’UTR (Fig. 1c). Fisher’s exact test further showed that DMPs were significantly enriched in distal promoter (TSS1500, *P* = 0.003), first exon (P = 0.008) and intergenic region (*P* = 0.004), while being depleted from gene bodies (*P* = 0.018), UTRs (5’UTR, *P* = 0.002; 3’UTR *P*= 0.031) and proximal promoter (TSS200, *P* = 0.024) (Fig. 1c). DMPs were not significantly enriched in FANTOM 5-annotated enhancers. Enrichment analysis according to CGI-related features revealed an enrichment of DMPs in shores (*P* = 6.04 × 10^−07^) and to a lesser extent in CGI (*P* = 0.001), along with depletion in open seas (*P* = 2.03 × 10^−05^) (Fig. 1d).

Altogether, these data imply noticeable DNA methylation changes in glial cells of MS patients compared to controls occurring in regulatory segments of genes.

### DNA methylation changes affect genes involved in cytoskeleton, motility, signaling and metabolism

To gain insight into the biological relevance of the DNA methylation changes in glial cells of MS patients, we conducted a Gene Set Analysis of genes harboring DMPs (*P*_adj_ < 0.05, 687 genes). Clustering of Gene Ontology (GO) terms indicated alteration of genes linked to six main functions: cytoskeleton organization, cell signaling, molecule transports, neurogenesis, cell motility and metabolic processes (Fig. 2a, Supplementary Table 4). Accordingly, altered genes form a biologically interconnected gene network (interaction enrichment *P*-value = 1.9 × 10^−03^) with the core DMP-genes involved in the GO categories (n =254 genes) presented in Figure 2b. More specifically, as listed in Table 3, DMP-genes implicated in cellular motility encode cell-adhesion molecules, such as cadherin and integrin, together with players of chemotaxis and extracellular matrix (ECM) remodelling (e.g. collagen and hyaluronan dynamics). Cytoskeleton dynamics genes are further represented by cytoskeleton-associated proteins (e.g. RhoGTPases) as well as vesicle-mediated transport linked to endocytosis, exocytosis and intracellular vesicle trafficking. The most represented intracellular signaling pathways are connected to Wnt/β-catenin and TGF-β/SMAD signaling followed by inflammation-mediated signaling pathways. Neurogenic signaling processes are for example reflected by molecules involved in signaling through glutamate and GABA, along with players of axo-glial processes, including myelination. Metabolic processes encompass differential methylation at genes encoding proteins involved in mitochondria integrity and oxidative stress, nutrient homeostasis, molecule degradation and ion transport (predominantly potassium channels). Transcriptional regulation comprises a plethora of DNA-binding transcription factors involved in nervous and immune cell fate, proliferation and cell arrest as well as DNA repair and chromatin regulation. Differential methylation also implicated genes involved in RNA synthesis including ribosomal and viral transcription. GO analysis of genes harboring DMP conditioned to their genomic location showed that enrichment of these processes arise from DMPs located in most gene segments (Supplementary Fig. 3).

**Figure 2.**
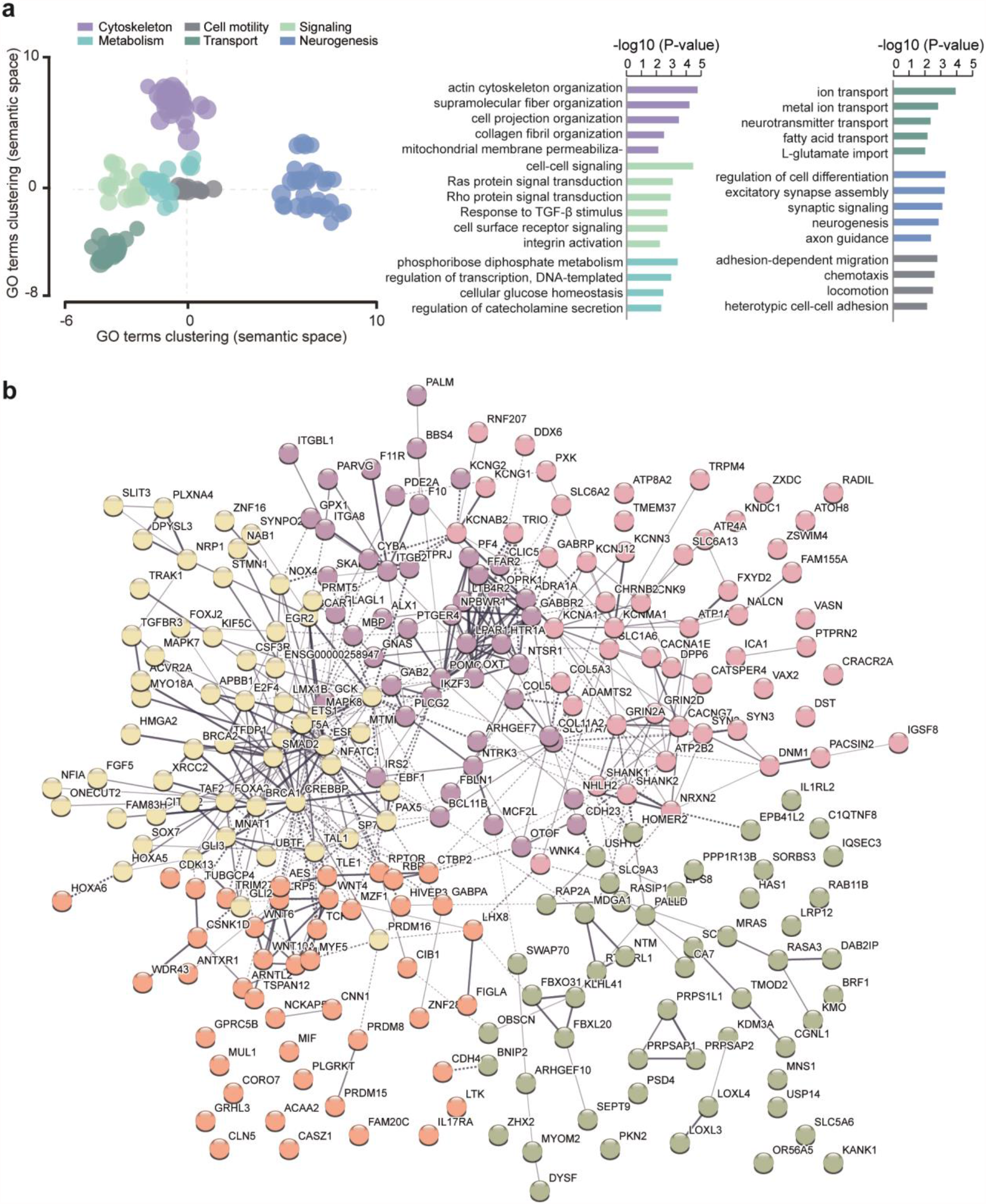
Functional annotation of glial DNA methylation changes associated with Multiple Sclerosis **a**. Scattered plot showing clusters of gene ontology (GO) terms associated with differentially methylated genes (DMP with *P*_adj_ < 0.05) in MS patients compared to NNC. *Biological Processes* GO terms were obtained using over-representation analysis and clusters were visualized in two-dimensional space (assigning *x* and *y* coordinates to each term) by applying multidimensional scaling of the matrix of GO terms according to semantic similarity, using REVIGO. The circle size represents −log10 (*P-*value), with small to big diameters ranging from 2.07 (*P* = 8 × 10^−03^) to 5.44 (*P* = 3 × 10^−06^). **b**. Top GO terms associated to different categories of *Biological processes* **c**. Representation of the genes containing DMPs (*P*_adj_ < 0.05) involved in the *Biological processes* using STRING network analysis. Grey gradient indicated the strength of data support (darker grey representing stronger evidence, dotted line showing lower level of evidence, i.e. combined interaction score 0.4-0.6). Colors represent different clusters (kmeans clustering set at 5 clusters). Full GO data are presented in Supplementary Table 4.

**Table 3.**
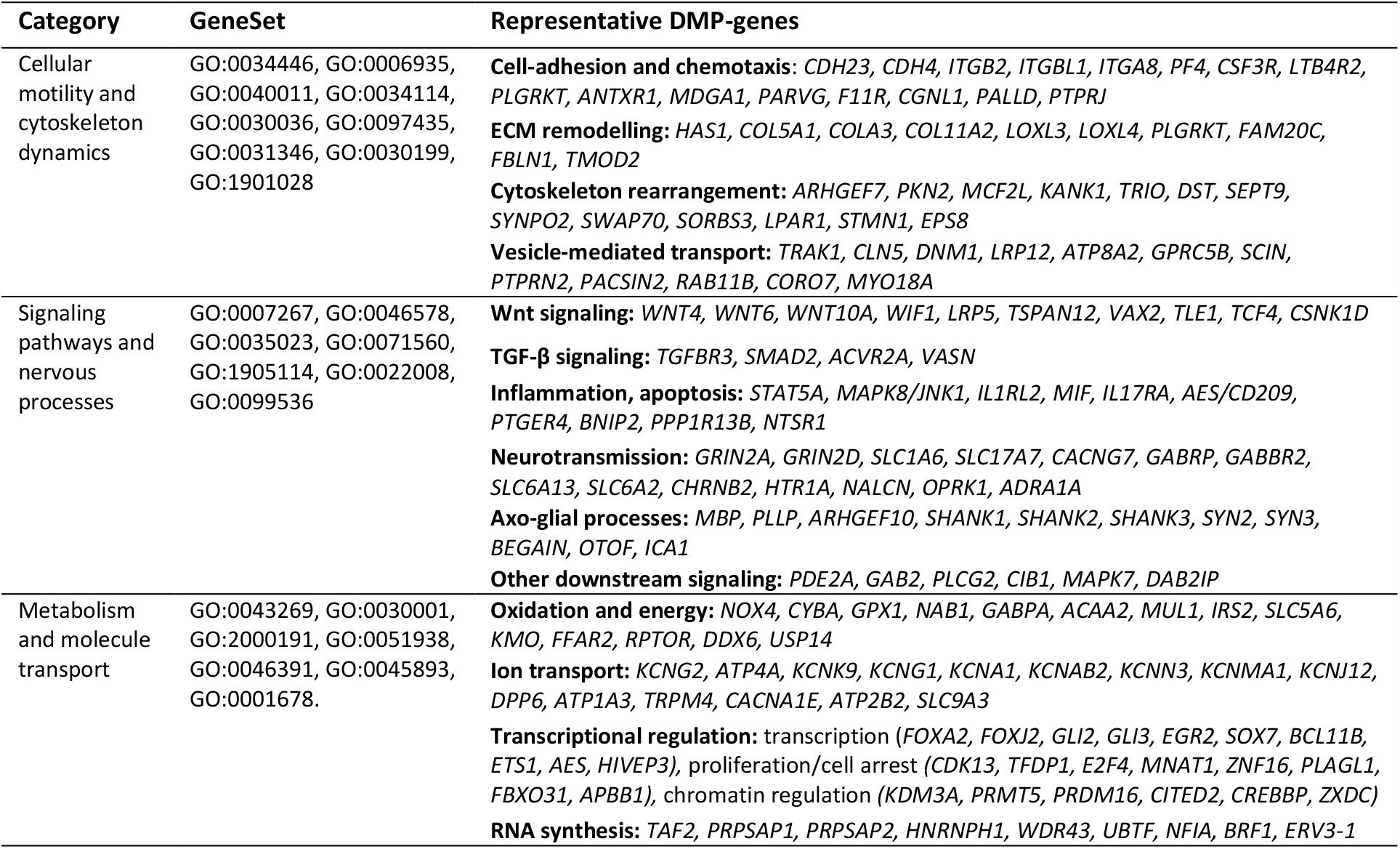
Gene ontology analysis of differentially methylated genes (DMP with *P*_adj_ < 0.05) associated with Multiple Sclerosis in glial nuclei

Functional annotation of DMRs confirmed GO analysis of DMPs with enrichment of ECM and migration (e.g. *ITGB2, MANBA, NEU4, SCIN, STMN1*), TGF-β and Wnt signaling pathways (*GREM2, SMAD2, TLE1*) and metabolism (*NOX5, CRYZ, CYP1A1, LDHAL6A*) among others (Supplementary Table 4). Examples of DMR genes implicated in these processes are illustrated in Figure 3. They encode cell surface integrin subunit (*ITGB2*) involved in adhesion, migration and phagocytosis, among others, mitochondrial cytochrome P450-mediated enzymes (*GSTM5*), developmental transcription factor antagonizing Wnt signaling (*VAX2)*, key players in involved in neurogenesis and iron homeostasis (*SEZ6L2* and *BOLA2*, respectively), insertase to the ER membrane (*WRB*), mediator of necroptosis (*MLKL*) and IFN-induced dynamin-like GTPase (*MX2*).

**Figure 3.**
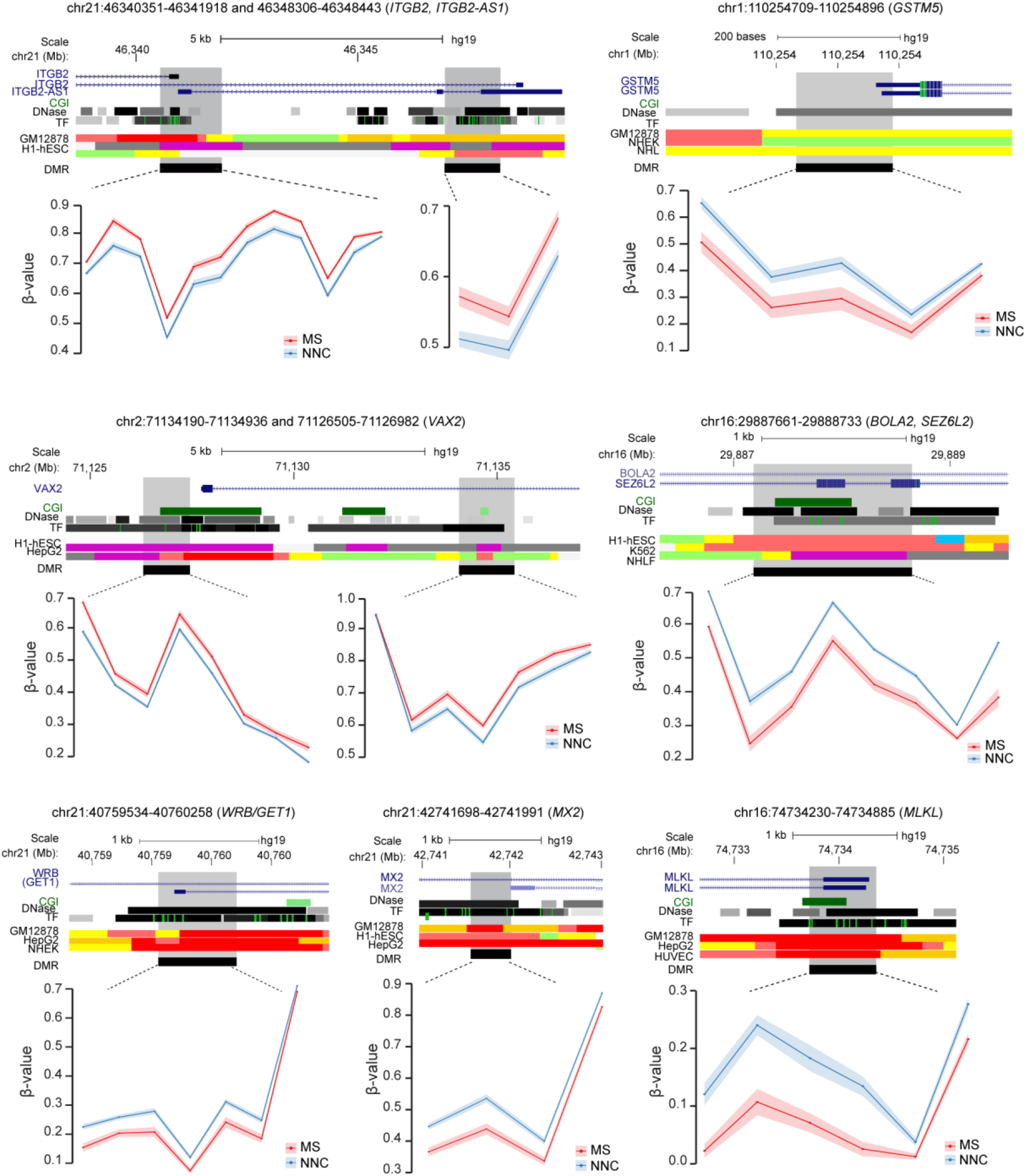
Differentially methylated regions in glial cells of Multiple Sclerosis patients. Plots illustrating genes associated with differentially methylated regions (DMRs) identified in glial nuclei from Multiple Sclerosis (MS) cases compared to non-neurological controls (NNC). The hg19 ideogram illustrating chromosome and cytoband information, the complete gene structure of the locus (blue track), CpG Island (CGI) (green track), regulatory properties exemplified in cell lines from chromatin state segmentation by hidden Markov model from ENCODE/Broad and DMR (black) location are shown. Methylation (β-values) of single consecutive CpGs within the DMR for cases and controls is depicted in red and blue, respectively, with connecting lines indicating mean methylation ± SEM.

### DNA methylation changes are associated with gene expression differences in MS glial cells

We sought to further explore the putative functional impact of the identified methylation changes by examining expression of the corresponding genes from published RNA-sequencing data conducted in post-mortem brain MS and controls samples. Comparison of differentially methylated genes with transcripts detected in RNA-seq data of bulk WM transcriptome^33^ showed that a minor fraction (∼14%, 80/561) displayed significant transcriptional differences, as reported in the original analysis with *P* < 0.05 (Fig. 4, Supplementary Table 2). The majority of them (51/80) were found downregulated in MS NAWM compared to control WM, this association being higher than expected for genes containing DMPs within gene body (Chi-square test *P* = 0.019) (Fig. 4). Overall, the transcriptionally dysregulated DMP-genes encode proteins primarily involved in adhesion and migration of nervous cells (e.g. *SLIT3, DAB2IP, GLI3, NRP1, CDH4* genes) as well as cytoskeleton remodeling and vesicle trafficking (e.g. *ATP8A2, TMOD2, OBSCN, TRAK1, KIF5C, PARVG* genes) and inflammatory response (e.g. *DAB2IP, ITGB2, CYBA, LILRA4* genes). Comparison of DMP-genes with single-cell RNA-seq findings from two studies profiling MS-lesions compared to NNC brain tissue^13,14^ indicated very minor overlap between DMP-genes (7%, 48/687) and differentially expressed genes in a given cell type of MS patients compared to the corresponding one of NNC individuals (Supplementary Table 2). Some of the differentially methylated and expressed genes found in oligodendrocytes (*RBFOX1, NTM, PPP1R12B* genes), microglia (*GPX1, HIVEP3, KCNMA1, FYB, ARHGAP22* genes) or astrocytes (*PRDM16, GLI3, WIF1* genes) from single-cell studies overlapped with dysregulated genes identified in bulk NAWM brain tissue of MS patients compared to WM of NNC individuals (Supplementary Table 2).

**Figure 4.**
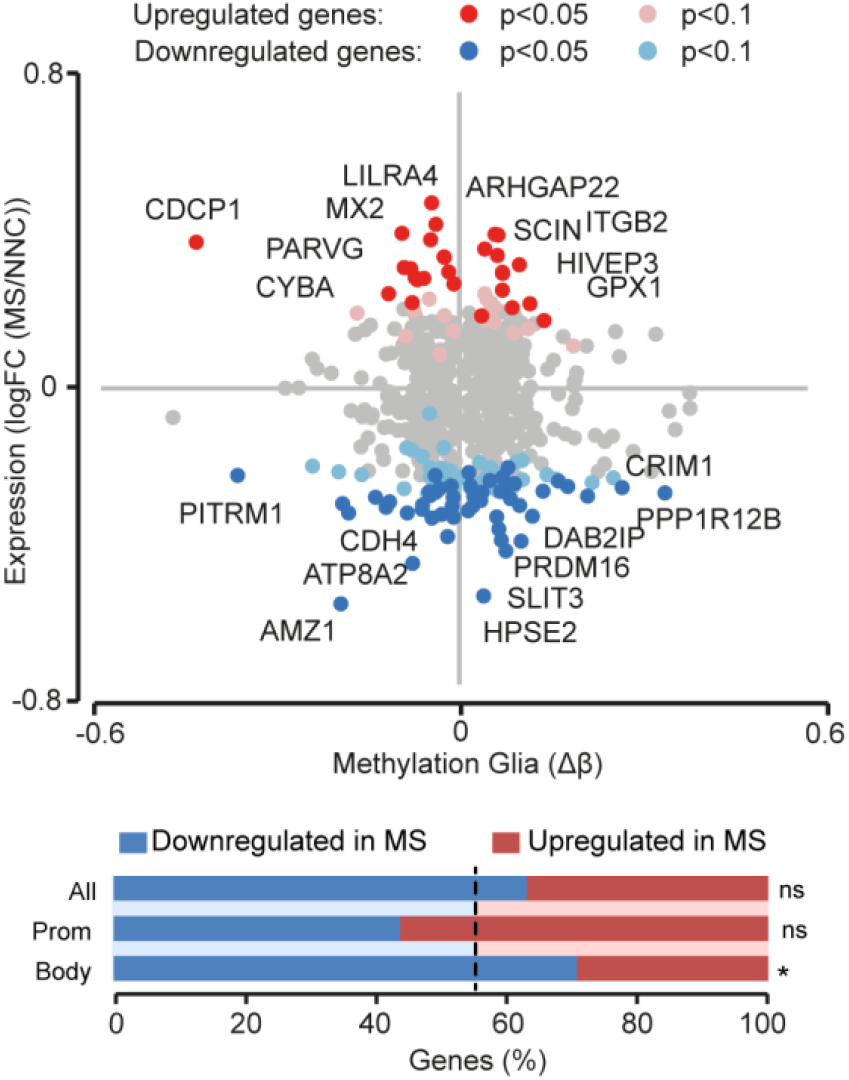
Association of DNA methylation changes with gene expression in glia of Multiple Sclerosis patients. Scatterplot illustrating association of DMPs (*P*_adj_ < 0.05) methylation values (Δβ) in glial nuclei samples with gene expression data (RNA-seq) reported in bulk NAWM of Multiple Sclerosis (MS) patients compared to WM of non-neurological controls (NNC)^33^. Red and blue colors indicate upregulation and downregulation, respectively, in MS versus NNC bulk brain tissue. The barplot represents the proportions (percentage) of upregulated (red) and downregulated (blue) genes for all gene-annotated DMPs and DMPs located in promoter (TSS1500, TSS200) or gene body. The dotted line indicating the expected proportion *\* P* < 0.05 generated with the Chi-square test.

These findings jointly suggest that methylation changes identified in glial nuclei of MS patients could, at least partly, be associated with transcriptional differences.

### DNA methylation changes affect shared and distinct pathways among glial cells

DNA methylation changes detected in glial cells likely reflect molecular changes occurring in several glial cell types. To further disentangle their possible contribution in MS brain, we assigned these alterations to specific glial cell types based on their constitutive expression in the healthy human brain^47,48^ (Supplementary Fig. 4, Supplementary Table 5). We explored the functional implication of epigenetic dysregulation at DMP-genes (*P*_adj_ < 0.05) assigned to astrocytes (n=309 genes), microglia (n=153 genes), and oligodendroyctes (n=196 genes) using GO analysis. Overall, GO terms clustered into distinct groups showing both common and distinct functions between glial cell types (Fig. 5a, Supplementary Table 4). MS-associated enrichment of biological functions related to cytoskeleton dynamics and ECM organization was shared by all glial cell types (Fig. 5b). Both microglia and astrocytes manifested enrichment of terms linked to adhesion and differentiation while formation of cellular projection and neurogenesis could be associated to oligodendrocytes and astrocytes (Fig. 5b).

**Figure 5.**
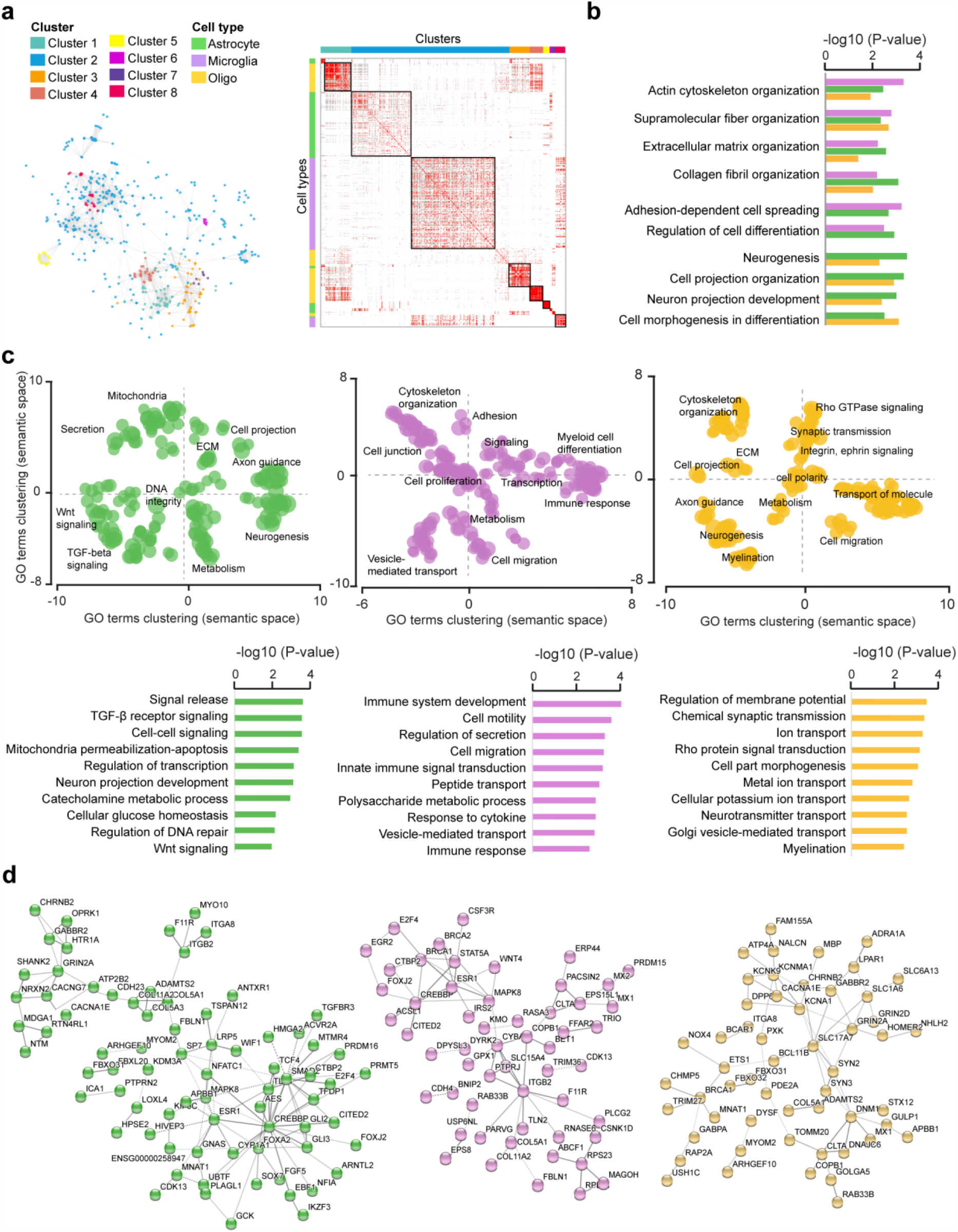
Cell type-specific functional association of DNA methylation changes in glial cells of Multiple Sclerosis patients. **a**. Heatmap of significant GO terms related to Biological Processes (min 3 molecules) generated from genes that associate with cell type-annotated DMPs. GO terms were grouped into clusters using the relative risk (RR) between different pathways, which was calculated based on the number of overlapping genes per pathway^46^. The left panel reflects relative distance between GO terms and clusters. **b**. Significantly enriched *Biological processes* shared by at least 2 out of 3 cell types. **c**. Scattered plot showing clusters of gene ontology (GO) terms associated with differentially methylated genes (DMP with *P*_adj_ < 0.05) in MS patients compared to NNC, annotated as expressed in astrocytes (left), microglial cells (middle) and oligodendrocyte (right). *Biological Processes* GO terms were obtained using over-representation analysis and clusters were visualized in two-dimensional space (assigning *x* and *y* coordinates to each term) by applying multidimensional scaling of the matrix of GO terms according to semantic similarity, using REVIGO. The circle size represents −log10 (*P-*value), with small to big diameters ranging from 1.32 (*P* = 0.04) to 4.2 (*P* = 6.5 × 10^−05^). **d**. Top *Biological processes* associated with differentially methylated genes (DMP with P_adj_ < 0.05) in MS patients compared to NNC, annotated as expressed in astrocytes (left), microglial cells (middle) and oligodendrocyte (right). **e**. Representation of the cell type-restricted genes containing DMPs (*P*_adj_ < 0.05) involved in the top 50 *Biological processes* from GO analysis in astrocytes (green), microglia (purple) and oligodendrocytes (orange), using STRING network analysis. Grey gradient indicated the strength of data support (darker grey representing stronger evidence, dotted line showing lower level of evidence, i.e. combined interaction score 0.4-0.6). Full GO data are presented in Supplementary Table 4. Astrocytes, microglial cells and oligodendrocyte are depicted in green, purple and orange, respectively.

In addition to these shared cellular functions, distinct signatures could be observed for each cell type-assigned DMP-gene sets (Fig. 5c). Astrocyte-annotated DMP-genes were predominantly implicated in intracellular signaling such as TGF-β (*TGFBR3, SMAD2, ITGA8, VASN*) and Wnt (*GLI3, WIF1, SOX, TSPAN12, AES, TLE1*) signaling pathways, neurotransmission (reflected by several neurotransmitter receptors), along with mitochondria damage (*ACAA2, MUL1, MAPK8*/*JNK1, PPP1R13B*), DNA repair (*FAM168A, BPHL, APBB1, MGMT, RTEL1*) and regulation of transcription (represented by multiple transcriptional regulators). Microglial DMP-genes showed an overrepresentation of innate immune-related functions such as inflammatory response (*IL1RL2, FFAR2, SLC15A4, MAPK8*/*JNK1, PTGER4, SWAP70, IL17RA, PLCG2*), defense response (*LILRA4, FCGRT, MX1*/*2, RNASE6, ALPK1, CDK13*), oxidative stress (*CYBA, GPX1*), cell motility (*CDH4, CSF3R, ITGB2, HAS1, COL5A1, USP14*), signaling (*CITED2, CSNK1D*). The analysis also underscored genes implicated in vesicle-mediated transport involved in endocytosis (*CLTA, FCGRT, SCIN, PACSIN2, USP6NL, RIN2, RUFY1*), ER-Golgi trafficking and autophagy (*BET1, COPB1, RAB33B*) as well as co-and cargo-transport (*CNST, SLC15A4*). Functions enriched in DMP-genes assigned to oligodendrocytes were connected to synaptic transmission (*PXK, USP14, ADRA1A, SYN3, GRIN2A*/*2D, CACNA1E*), potassium and sodium transport (*DPP6, KCNG1, KCNA1, NALCN, FAM155A, PDE2A*), cytoskeletal rearrangement (*STMN1, LPAR1, RAP2A, ARHGEF7, KANK1, KIF5C*), signaling adaptor protein (*GAB2, APBB1*) and myelination (*ARHGEF10, MBP, NAB1, PLLP*). Examples of gene networks associated to GO terms in each cell type are illustrated in Figure 5d. GO analysis of the cell type-assigned DMP-genes according to their gene location indicated that the majority of the enriched biological processes reflect methylation changes occurring in the gene body and further uncovered specific functions that are more enriched in promoter-occupying DMPs (Supplementary Fig. 5, Supplementary Table 4).

Reactome clustering of biological interactions confirmed the GO findings and further delineated specific pathways within each biological process (Supplementary Fig. 6, Supplementary Table 4). Accordingly, Wnt signaling found enriched in astrocyte-assigned genes arise primarily from the *Repression of Wnt target genes* set. Terms related to neurotransmission characterizing astrocyte- and oligodendrocyte-assigned DMP-genes were involved, among others, in protein-protein interaction at synapses pathways such as *Neurexins and Neurogilins*. Transcriptional processes found enriched in DMPs-gene assigned to all cell types referred to *FOXO-mediated transcription*, while astrocytic and microglial DMP-genes were additionally involved in *Transcriptional regulation by AP-2 and TP53*.

These findings collectively support alterations of shared processes prevailing in all cell types, such as cytoskeleton organization, as well as putative distinct cell type-specific functions converging to Wnt/ TGF-β signaling in astrocytes, cellular motility and innate immunity in microglia and ion transport and neuromodulation in oligodendrocytes.

## Discussion

We exploited the stable nature of DNA methylation, informing on the genome activity in post-mortem material, to investigate the still elusive molecular changes occurring in NAWM glial cells of MS patients in comparison to WM glial cells of NNC individuals. We found DNA methylation changes at genes involved in cellular motility, cytoskeleton rearrangement, cell-to-cell and intracellular signaling, such as Wnt and TGF-β signaling, neuromodulation and neuroinflammation, among others, a fraction of these genes were previously shown to be dysregulated in the brain of MS patients. Our findings strongly suggest that NAWM glial cells in MS are highly altered in the absence of lesional insult, collectively exhibiting a multicellular response to diffuse inflammation. Whether the observed changes reflect pre-lesional processes or tissue reaction to adjoining inflammatory damage remains to be elucidated.

Glial cells of MS patients exhibited epigenetic alterations affecting several genes implicated in cellular and intracellular motility, from substrate adhesion molecules and cell-cell junction to cytoskeleton dynamics and ECM remodeling. Differences in cellular migration and cytoskeleton dynamics have been described in developing or healthy CNS tissue and in the context of focal insult, such as MS demyelinating plaques^49,50^. The identification of such alterations in our cohort, composed of NAWM tissue, supports the occurrence of abnormalities in the unaffected areas as well, as previously suggested in NAWM bulk tissue and neurons^33,34^ and might reflect tapering gradient of reactive gliosis or focal event prior to injury. In line with this, active microglia assume enhanced agility and rapidly converge to sites of damage, directional migration following a chemoattractant gradient of soluble molecules released by damaged cells and intertwined astrocytes^10,51^. Accordingly, dysfunctional astrocytes present with retracted processes and loss of cell-cell junctions, and ablation of reactive astrocytes hinders the recruitment of microglia to demyelinating lesions^24,52^. Such inhibition has been shown to impair debris clearance, oligodendrocyte maturation and remyelination^53^. The latter process also mobilizes intense cytoskeleton dynamics involved in the formation of growth cones and amyloid-like myelin-enriched protrusions ensheathing and enwrapping denuded axons^54^.

Alteration of genes connected to cell-to-cell and intracellular signaling through Wnt and TGF-β families further underscore a coordinated response in the NAWM of MS patients due to to diffuse damage and confirmed previous observation of aberrant Wnt and TGF-β activity in the MS brain^27,55,56^. A pivotal role of TGF signaling in experimental autoimmune, neuroinflammatory or demyelinating diseases is already highly recognized^57^. Notably, the differentially methylated genes of Wnt/β-catenin pathways operate at multiple levels of the signaling cascade, from complex formation between extracellular Wnt ligand (*WNT4, WNT6, WNT10A*), Wnt ligand antagonist (*WIF1*) and receptors (*LRP5*), to subsequent β-catenin stabilization (*TSPAN12)* and activation of TCF/LEF-mediated transcriptional program (*TLE1, TCF4, VAX2*). Dysfunctional Wnt pathways in MS-like animal models has further shown to enhance astrogliosis, hinder remyelination, and impair cell migration to the demyelinated lesions^55,58–60^.

Interestingly, glial cells of the MS-NAWM displayed epigenetic alterations of molecules involved in neuronal integrity and plasticity, compared to WM glial cells of controls. Methylation changes affected genes encoding synapse scaffolding and clustering proteins, such as postsynaptic density proteins (PSD), synapsins and neurexin, and can be exemplified by the robust hypermethylation (DMP with Δβ > 0.15), spanning over a DMR at exon 2 of the PSD95-associated *BEGAIN* gene. Hypermethylation of the very same region has been identified in bulk brain tissue and glia of patients affected by neuropsychiatric disorders and inversely correlated with expression changes^61^. More generally, genetic aberrations of many of the differentially methylated genes in our study, i.e. *TRIO, NRXN2, SYN2, CACNA1E, DNM1* and *RAB11B*, have been associated with movement disorders, cognitive and visual impairments and brain abnormalities (e.g. atrophy, white matter apoplasia)^62^. Methylation changes at genes encoding modulators of synaptic transmission, in particular potassium/calcium transport and glutamate transmission further reinforces the role of glial cells in supporting neuronal functions in general. Accordingly, the glial control of neuronal activity strongly relies on extracellular potassium control, i.e. both astrocytes and oligodendrocytes respond to neuronal discharge and reciprocally coordinate neuronal excitability by buffering/dispersing extracellular potassium^63^. They operate via potassium channels (illustrated by differentially methylated genes encoding voltage-gated, calcium-activated and inwardly rectifying K^+^ channels in our cohort) and astrocyte-oligodendrocyte junctional coupling embedded in a panglial syncytium^64–66^. Additionally, astrocytes govern neuromodulatory networks and promote excitatory signaling via sheathing of a vast number of tripartite synapses, expression of neuromodulatory receptors, glutamate uptake and secretion of gliotransmitters^67,68^. While this finding reflects the dynamic features of glial cells of the NAWM, the circumstances underlying such alterations in the seemingly unaffected tissue, i.e. as a cause or consequence of brain disconnectivity, warrant further investigation.

Because epigenetic marks are highly tissue- and cell type-specific, especially in the CNS^69,70^, the analysis of the NeuN-negative fraction sorted from brain tissue could undeniably be biased by cellular heterogeneity. To mitigate the impact of such potential confounder, we analyzed sorted nuclei from the WM exclusively and further applied reference-free deconvolution accounting for varying proportions of glial cell types. However, one cannot exclude the possibility of missed signals and, whether the identified changes reflect large differences in one cell type and/or shared variations in several glial cell types remain to be elucidated. Overall, our approach likely captures the molecular signature of interwoven processes, as reported at the transcriptomic level as well^27^. We attempted to delineate common and cell type-specific process by integrating single-cell transcriptomic data. Cell type-specific functional annotation unambiguously attributed abnormalities of cytoskeleton dynamics and ECM organization to all glial cell types. GO analyses implied a pervasive role of oligodendrocytes and astrocytes in supporting neuronal integrity via cell-cell signaling and neuromodulation, among others and unveiled more specific functions in microglia, converging to cell migration, vesicle-mediated transport, RNA metabolism and neuroinflammation such as antiviral processes. This can be exemplified by large (Δβ> 0.15) and wide (DMR) hypermethylation at *ERV3-1* gene, encoding human endogenous retrovirus (HERV)-R element. HERVs have gained large interest in neurodegenerative diseases in general and in MS in particular, notably as disease markers and putative targets for MS therapy^71–73^, with a still unclear role of HERV-R^74,75^. Whether changes at *ERV1-3* locus in glial cells of MS patients reflect the physiological role of HERV-R (*per se* or *via* regulation of neighboring genes) as reported^76^ or rather a pathogenic event, as described for other HERVs remain to be ascertained.

In conclusion, DNA methylation analysis of glial nuclei sorted from post-mortem WM of MS patients and controls unraveled molecular changes affecting motile, signaling and neuromodulatory properties of NAWM glial cells. These alterations likely compile multiple discrete focal insults impinging the CNS in absence/prior to lesion or alternatively reflect compensatory mechanisms circumventing the gradual and global deterioration of the neural circuitry. Overall, our findings portray the NAWM as a highly dynamic tissue in response to the persistent inflammation endured by the CNS in MS patients. The identification of epigenetic abnormalities, which are modifiable by nature, affecting theses processes provides additional evidence in favor of therapeutic strategies aiming at rehabilitating the functional capacities of the CNS in progressive MS patients by targeting CNS resident cells.

## Supporting information

Supplementary Table 1_revised

## Data Availability

The EPIC data that support the findings of this study are available in Gene Expression Omnibus (GEO) database under the accession number GSE166207.

## Contributors

MJ, LK and MN conceived and designed the study. MJ supervised the study. LK and RC performed sample isolation and FACS sorting. MPK conducted DNA extraction. EE carried out DNA methylation analysis and MN and DGC contributed to the analytical pipeline with input. LK conducted additional analyses. LB aided in accessing the samples. LK wrote the manuscript with assistance from all authors. All authors read and approved the manuscript.

## Acknowledgments

We are grateful to A. van Vollenhoven for flow cytometry processing (Center for Molecular Medicine and Karolinska Institutet core facility). We acknowledge GenomeScan/ServiceXS (Leiden, The Netherlands) for processing Illumina EPIC arrays. We thank the Multiple Sclerosis and Parkinson’s Tissue Bank (Imperial College London) for provision of brain tissue samples. Computations were performed on resources provided by SNIC through Uppsala Multidisciplinary Center for Advanced Computational Science (UPPMAX).

## References

1. Frischer, J.M. et al. Clinical and pathological insights into the dynamic nature of the white matter multiple sclerosis plaque. Ann Neurol 78, 710–21 (2015).

2. Kornek, B. et al. Multiple sclerosis and chronic autoimmune encephalomyelitis: a comparative quantitative study of axonal injury in active, inactive, and remyelinated lesions. Am J Pathol 157, 267–76 (2000).

3. Frischer, J.M. et al. The relation between inflammation and neurodegeneration in multiple sclerosis brains. Brain 132, 1175–89 (2009).

4. Nikic, I. et al. A reversible form of axon damage in experimental autoimmune encephalomyelitis and multiple sclerosis. Nat Med 17, 495–9 (2011).

5. Bodini, B. et al. Individual Mapping of Innate Immune Cell Activation Is a Candidate Marker of Patient-Specific Trajectories of Worsening Disability in Multiple Sclerosis. J Nucl Med 61, 1043–1049 (2020).

6. Yeung, M.S.Y. et al. Dynamics of oligodendrocyte generation in multiple sclerosis. Nature 566, 538–542 (2019).

7. Goldschmidt, T., Antel, J., Konig, F.B., Bruck, W. & Kuhlmann, T. Remyelination capacity of the MS brain decreases with disease chronicity. Neurology 72, 1914–21 (2009).

8. Ransohoff, R.M. How neuroinflammation contributes to neurodegeneration. Science 353, 777–83 (2016).

9. von Bartheld, C.S., Bahney, J. & Herculano-Houzel, S. The search for true numbers of neurons and glial cells in the human brain: A review of 150 years of cell counting. J Comp Neurol 524, 3865–3895 (2016).

10. Nimmerjahn, A., Kirchhoff, F. & Helmchen, F. Resting microglial cells are highly dynamic surveillants of brain parenchyma in vivo. Science 308, 1314–8 (2005).

11. van der Poel, M. et al. Transcriptional profiling of human microglia reveals grey-white matter heterogeneity and multiple sclerosis-associated changes. Nat Commun 10, 1139 (2019).

12. Liddelow, S.A. et al. Neurotoxic reactive astrocytes are induced by activated microglia. Nature 541, 481–487 (2017).

13. Schirmer, L. et al. Neuronal vulnerability and multilineage diversity in multiple sclerosis. Nature 573, 75–82 (2019).

14. Jakel, S. et al. Altered human oligodendrocyte heterogeneity in multiple sclerosis. Nature 566, 543–547 (2019).

15. Masuda, T. et al. Spatial and temporal heterogeneity of mouse and human microglia at single-cell resolution. Nature 566, 388–392 (2019).

16. Kutzelnigg, A. et al. Cortical demyelination and diffuse white matter injury in multiple sclerosis. Brain 128, 2705–12 (2005).

17. de Groot, M. et al. Changes in normal-appearing white matter precede development of white matter lesions. Stroke 44, 1037–42 (2013).

18. Gallego-Delgado, P. et al. Neuroinflammation in the normal-appearing white matter (NAWM) of the multiple sclerosis brain causes abnormalities at the nodes of Ranvier. PLoS Biol 18, e3001008 (2020).

19. Moll, N.M. et al. Multiple sclerosis normal-appearing white matter: pathology-imaging correlations. Ann Neurol 70, 764–73 (2011).

20. Barnett, M.H. & Prineas, J.W. Relapsing and remitting multiple sclerosis: pathology of the newly forming lesion. Ann Neurol 55, 458–68 (2004).

21. Henderson, A.P., Barnett, M.H., Parratt, J.D. & Prineas, J.W. Multiple sclerosis: distribution of inflammatory cells in newly forming lesions. Ann Neurol 66, 739–53 (2009).

22. van Horssen, J. et al. Clusters of activated microglia in normal-appearing white matter show signs of innate immune activation. J Neuroinflammation 9, 156 (2012).

23. Burm, S.M. et al. Expression of IL-1beta in rhesus EAE and MS lesions is mainly induced in the CNS itself. J Neuroinflammation 13, 138 (2016).

24. Sharma, R. et al. Inflammation induced by innate immunity in the central nervous system leads to primary astrocyte dysfunction followed by demyelination. Acta Neuropathol 120, 223–36 (2010).

25. Luchicchi, A. et al. Axon-Myelin Unit Blistering as Early Event in MS Normal Appearing White Matter. Ann Neurol (2021).

26. Werring, D.J. et al. The pathogenesis of lesions and normal-appearing white matter changes in multiple sclerosis: a serial diffusion MRI study. Brain 123 (Pt 8), 1667–76 (2000).

27. Elkjaer, M.L. et al. Molecular signature of different lesion types in the brain white matter of patients with progressive multiple sclerosis. Acta Neuropathol Commun 7, 205 (2019).

28. Ouellette, R. et al. Validation of Rapid Magnetic Resonance Myelin Imaging in Multiple Sclerosis. Ann Neurol 87, 710–724 (2020).

29. Kiljan, S. et al. Cortical axonal loss is associated with both gray matter demyelination and white matter tract pathology in progressive multiple sclerosis: Evidence from a combined MRI-histopathology study. Mult Scler 27, 380–390 (2021).

30. Kular, L. & Jagodic, M. Epigenetic insights into multiple sclerosis disease progression. J Intern Med 288, 82–102 (2020).

31. Neri, F. et al. Intragenic DNA methylation prevents spurious transcription initiation. Nature 543, 72–77 (2017).

32. Chomyk, A.M. et al. DNA methylation in demyelinated multiple sclerosis hippocampus. Sci Rep 7, 8696 (2017).

33. Huynh, J.L. et al. Epigenome-wide differences in pathology-free regions of multiple sclerosis-affected brains. Nat Neurosci 17, 121–30 (2014).

34. Kular, L. et al. Neuronal methylome reveals CREB-associated neuro-axonal impairment in multiple sclerosis. Clin Epigenetics 11, 86 (2019).

35. Morris, T.J. et al. ChAMP: 450k Chip Analysis Methylation Pipeline. Bioinformatics 30, 428–30 (2014).

36. Aryee, M.J. et al. Minfi: a flexible and comprehensive Bioconductor package for the analysis of Infinium DNA methylation microarrays. Bioinformatics 30, 1363–9 (2014).

37. Nordlund, J. et al. Genome-wide signatures of differential DNA methylation in pediatric acute lymphoblastic leukemia. Genome Biol 14, r105 (2013).

38. Chen, Y.A. et al. Discovery of cross-reactive probes and polymorphic CpGs in the Illumina Infinium HumanMethylation450 microarray. Epigenetics 8, 203–9 (2013).

39. Johnson, W.E., Li, C. & Rabinovic, A. Adjusting batch effects in microarray expression data using empirical Bayes methods. Biostatistics 8, 118–27 (2007).

40. Houseman, E.A., Molitor, J. & Marsit, C.J. Reference-free cell mixture adjustments in analysis of DNA methylation data. Bioinformatics 30, 1431–9 (2014).

41. Ritchie, M.E. et al. limma powers differential expression analyses for RNA-sequencing and microarray studies. Nucleic Acids Res 43, e47 (2015).

42. Du, P. et al. Comparison of Beta-value and M-value methods for quantifying methylation levels by microarray analysis. BMC Bioinformatics 11, 587 (2010).

43. Peters, T.J. et al. De novo identification of differentially methylated regions in the human genome. Epigenetics Chromatin 8, 6 (2015).

44. Zhang, B., Kirov, S. & Snoddy, J. WebGestalt: an integrated system for exploring gene sets in various biological contexts. Nucleic Acids Res 33, W741–8 (2005).

45. Supek, F., Bosnjak, M., Skunca, N. & Smuc, T. REVIGO summarizes and visualizes long lists of gene ontology terms. PLoS One 6, e21800 (2011).

46. Ewing, E., Planell-Picola, N., Jagodic, M. & Gomez-Cabrero, D. GeneSetCluster: a tool for summarizing and integrating gene-set analysis results. BMC Bioinformatics 21, 443 (2020).

47. Darmanis, S. et al. A survey of human brain transcriptome diversity at the single cell level. Proc Natl Acad Sci U S A 112, 7285–90 (2015).

48. Zhang, Y. et al. Purification and Characterization of Progenitor and Mature Human Astrocytes Reveals Transcriptional and Functional Differences with Mouse. Neuron 89, 37–53 (2016).

49. Frohman, E.M., Racke, M.K. & Raine, C.S. Multiple sclerosis--the plaque and its pathogenesis. N Engl J Med 354, 942–55 (2006).

50. Lucchinetti, C.F., Bruck, W., Rodriguez, M. & Lassmann, H. Distinct patterns of multiple sclerosis pathology indicates heterogeneity on pathogenesis. Brain Pathol 6, 259–74 (1996).

51. Davalos, D. et al. ATP mediates rapid microglial response to local brain injury in vivo. Nat Neurosci 8, 752–8 (2005).

52. Skripuletz, T. et al. Astrocytes regulate myelin clearance through recruitment of microglia during cuprizone-induced demyelination. Brain 136, 147–67 (2013).

53. Plemel, J.R., Manesh, S.B., Sparling, J.S. & Tetzlaff, W. Myelin inhibits oligodendroglial maturation and regulates oligodendrocytic transcription factor expression. Glia 61, 1471–87 (2013).

54. Thomason, E.J., Escalante, M., Osterhout, D.J. & Fuss, B. The oligodendrocyte growth cone and its actin cytoskeleton: A fundamental element for progenitor cell migration and CNS myelination. Glia 68, 1329–1346 (2020).

55. Fancy, S.P. et al. Dysregulation of the Wnt pathway inhibits timely myelination and remyelination in the mammalian CNS. Genes Dev 23, 1571–85 (2009).

56. Lee, H.K. et al. Daam2-PIP5K is a regulatory pathway for Wnt signaling and therapeutic target for remyelination in the CNS. Neuron 85, 1227–43 (2015).

57. Lund, H. et al. Fatal demyelinating disease is induced by monocyte-derived macrophages in the absence of TGF-beta signaling. Nat Immunol 19, 1–7 (2018).

58. Niu, J. et al. Aberrant oligodendroglial-vascular interactions disrupt the blood-brain barrier, triggering CNS inflammation. Nat Neurosci 22, 709–718 (2019).

59. Lengfeld, J.E. et al. Endothelial Wnt/beta-catenin signaling reduces immune cell infiltration in multiple sclerosis. Proc Natl Acad Sci U S A 114, E1168–E1177 (2017).

60. Robinson, K.F., Narasipura, S.D., Wallace, J., Ritz, E.M. & Al-Harthi, L. beta-Catenin and TCFs/LEF signaling discordantly regulate IL-6 expression in astrocytes. Cell Commun Signal 18, 93 (2020).

61. Nagy, C. et al. Astrocytic abnormalities and global DNA methylation patterns in depression and suicide. Mol Psychiatry 20, 320–8 (2015).

62. John, A., Ng-Cordell, E., Hanna, N., Brkic, D. & Baker, K. The neurodevelopmental spectrum of synaptic vesicle cycling disorders. J Neurochem (2020).

63. Beckner, M.E. A roadmap for potassium buffering/dispersion via the glial network of the CNS. Neurochem Int 136, 104727 (2020).

64. Menichella, D.M. et al. Genetic and physiological evidence that oligodendrocyte gap junctions contribute to spatial buffering of potassium released during neuronal activity. J Neurosci 26, 10984–91 (2006).

65. Brasko, C., Hawkins, V., De La Rocha, I.C. & Butt, A.M. Expression of Kir4.1 and Kir5.1 inwardly rectifying potassium channels in oligodendrocytes, the myelinating cells of the CNS. Brain Struct Funct 222, 41–59 (2017).

66. Battefeld, A., Klooster, J. & Kole, M.H. Myelinating satellite oligodendrocytes are integrated in a glial syncytium constraining neuronal high-frequency activity. Nat Commun 7, 11298 (2016).

67. Perea, G., Navarrete, M. & Araque, A. Tripartite synapses: astrocytes process and control synaptic information. Trends Neurosci 32, 421–31 (2009).

68. Bernardinelli, Y. et al. Activity-dependent structural plasticity of perisynaptic astrocytic domains promotes excitatory synapse stability. Curr Biol 24, 1679–88 (2014).

69. Sanchez-Mut, J.V. et al. Whole genome grey and white matter DNA methylation profiles in dorsolateral prefrontal cortex. Synapse 71(2017).

70. Davies, M.N. et al. Functional annotation of the human brain methylome identifies tissue-specific epigenetic variation across brain and blood. Genome Biol 13, R43 (2012).

71. Kornmann, G. & Curtin, F. Temelimab, an IgG4 Anti-Human Endogenous Retrovirus Monoclonal Antibody: An Early Development Safety Review. Drug Saf 43, 1287–1296 (2020).

72. Dolei, A., Ibba, G., Piu, C. & Serra, C. Expression of HERV Genes as Possible Biomarker and Target in Neurodegenerative Diseases. Int J Mol Sci 20(2019).

73. Gottle, P. et al. Rescuing the negative impact of human endogenous retrovirus envelope protein on oligodendroglial differentiation and myelination. Glia 67, 160–170 (2019).

74. Rasmussen, H.B., Heltberg, A., Lisby, G. & Clausen, J. Three allelic forms of the human endogenous retrovirus, ERV3, and their frequencies in multiple sclerosis patients and healthy individuals. Autoimmunity 23, 111–7 (1996).

75. Rasmussen, H.B. et al. Expression of endogenous retroviruses in blood mononuclear cells and brain tissue from multiple sclerosis patients. Mult Scler 1, 82–7 (1995).

76. Bustamante Rivera, Y.Y., Brutting, C., Schmidt, C., Volkmer, I. & Staege, M.S. Endogenous Retrovirus 3 - History, Physiology, and Pathology. Front Microbiol 8, 2691 (2017).

