## Supplementary Figures for "DNA methylation changes in glial cells of the normal-appearing white matter in Multiple Sclerosis patients"

#### Supplementary Tables description

**Supplementary Table 1.** Description of the cohort. MS, Multiple Sclerosis, NNC, non-neurological disease controls, F, female, M, male, NAWM, normal-appearing white matter, WM, white matter.

**Supplementary Table 2.** DNA methylation changes in glial cells of Multiple Sclerosis (MS) patients compared to non-neurological disease control (NNC) ($P_{adj} < 0.05$). Gene expression (RNA-sequencing) data in MS vs. controls bulk brain tissue (Huynh *et al*., 2014) or in single cells (Schirmer *et al*., 2019, Jäkel & Agrirre *et al*., 2018) are indicated for each identified gene.

**Supplementary Table 3.** Differentially methylated regions (DMR) in glial cells of Multiple Sclerosis (MS) patients compared to non-neurological disease control (NNC).

**Supplementary Table 4.** Gene ontology (GO) analyses. Each sheet lists data from GO analyses performed on all DMPs ($P_{adj} < 0.05$), microglia-, oligodendrocyte-, astrocyte-annotated DMPs ($P_{adj} < 0.05$) as well as GenesetCluster and Reactome analyses.

**Supplementary Table 5.** DMP-genes annotated according to constitutive expression in astrocytes, microglia and oligodendrocytes. Gene expression data from the healthy brain (Darmanis *et al*., 2015; Zhang *et al*., 2016). nd, not definable, na, not available.

#### Supplementary Figure 1

**a**

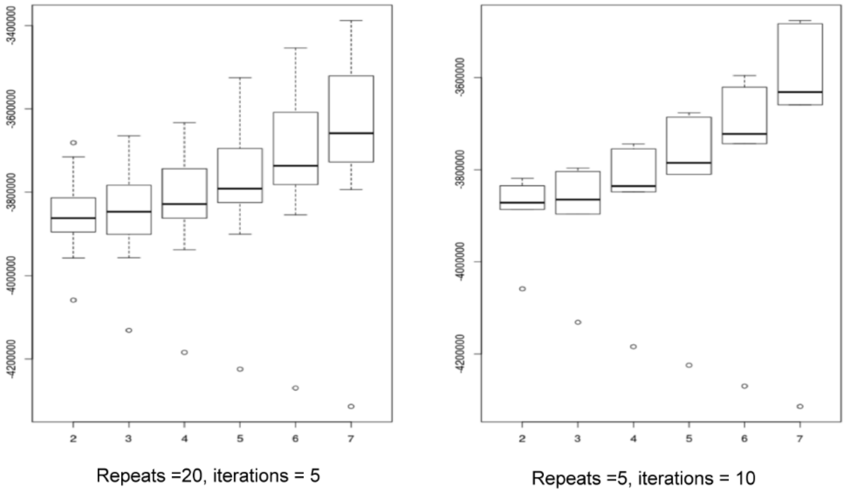


**b**

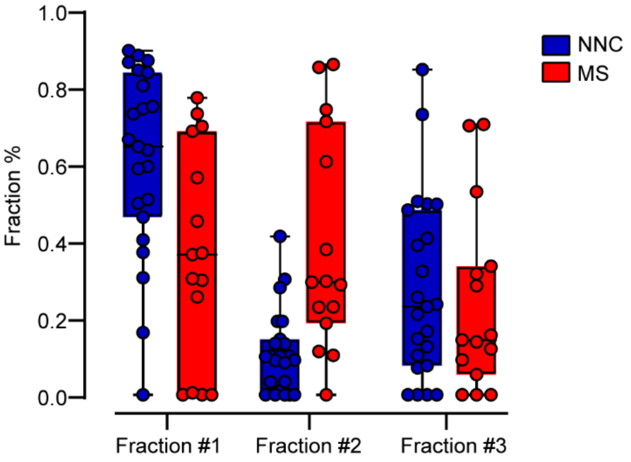


| | Mean NNC | SD NNC | Mean MS | SD MS | Pvalue |
|---|---|---|---|---|---|
| **Fraction #1** | 0.61 | 0.24 | 0.37 | 0.28 | 0.0102 |
| **Fraction #2** | 0.11 | 0.11 | 0.39 | 0.29 | 0.0020 |
| **Fraction #3** | 0.27 | 0.24 | 0.24 | 0.24 | 0.6552 |

**Supplementary Figure 1.** Deconvolution.

**a.** Deviance plotting from RefFreeEwas shows the accuracy of the model with the minimal amount of deviance around three different fractions using both more repeats (left) and more iterations (right) of the model. **b.** Plot and Table containing the mean and standard deviation (SD) of each fraction per MS and NNC samples with the independent samples T-test *P*-value.

#### Supplementary Figure 2

**a**

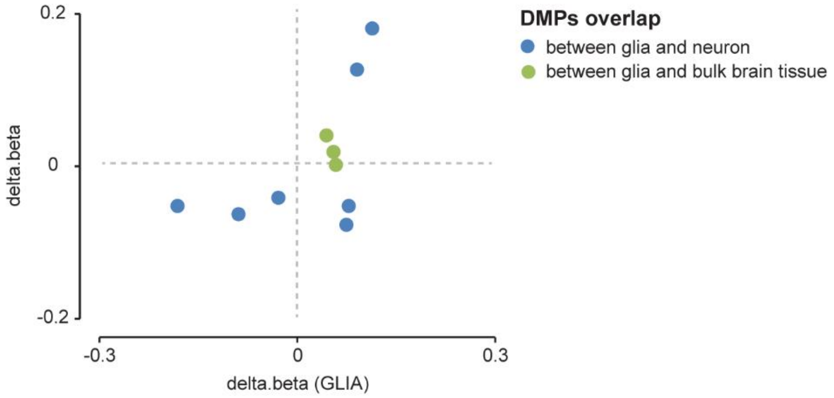


**b**

| Description | | | | | | Glia | | | | | Neurons | | |
|---|---|---|---|---|---|---|---|---|---|---|---|---|---|
| Probes | Chr. | Position | gene | feature | cgi | P.Val | adj.P.Val | Mean.MS | Mean.NNC | Δβ | Mean.MS | Mean.NNC | Δβ |
| cg02282631 | 5 | 42953543 | | IGR | shore | 1.1E-05 | 1.6E-02 | 0.209 | 0.188 | 0.079 | 0.117 | 0.145 | -0.028 |
| cg03839074 | 5 | 43000303 | | IGR | shore | 5.5E-05 | 3.9E-02 | 0.345 | 0.331 | 0.077 | 0.160 | 0.199 | -0.040 |
| cg04831505 | 12 | 72233240 | TBC1D15 | TSS1500 | shore | 1.3E-06 | 4.3E-03 | 0.471 | 0.373 | 0.116 | 0.413 | 0.325 | 0.088 |
| cg05241461 | 19 | 22816980 | ZNF492 | TSS200 | shore | 8.6E-05 | 4.9E-02 | 0.038 | 0.064 | -0.029 | 0.053 | 0.074 | -0.022 |
| cg13264183 | 3 | 126194992 | ZXDC | TSS1500 | island | 5.9E-06 | 1.1E-02 | 0.304 | 0.232 | 0.093 | 0.213 | 0.151 | 0.061 |
| cg16206678 | 6 | 30122152 | TRIM10 | Body | opensea | 2.1E-05 | 2.3E-02 | 0.703 | 0.762 | -0.090 | 0.771 | 0.804 | -0.033 |
| cg22992730 | 19 | 4784940 | | IGR | shore | 3.8E-05 | 3.3E-02 | 0.570 | 0.760 | -0.184 | 0.062 | 0.090 | -0.027 |

| Description | | | | | | Glia | | | | | Bulk tissue | | |
|---|---|---|---|---|---|---|---|---|---|---|---|---|---|
| Probes | Chr. | Position | gene | feature | cgi | P.Value | adj.P.Val | Mean.MS | Mean.NNC | Δβ | Mean.MS | Mean.NNC | Δβ |
| cg05930207 | 17 | 38518520 | GJD3 | 1stExon | shore | 5.6E-06 | 1.1E-02 | 0.760 | 0.714 | 0.060 | 0.683 | 0.684 | -0.001 |
| cg17383186 | 1 | 154127629 | NUP210L | TSS200 | opensea | 5.9E-06 | 1.1E-02 | 0.289 | 0.278 | 0.056 | 0.290 | 0.272 | 0.018 |
| cg05245094 | 16 | 85669572 | KIAA0182 | 5'UTR | shore | 6.0E-05 | 4.1E-02 | 0.836 | 0.805 | 0.044 | 0.784 | 0.776 | 0.008 |

**Supplementary Figure 2.** Overlap of DMPs with bulk brain tissue and neuron analyses.

**a.** Dot plot correlating the effect size of differentially methylated CpGs overlapping between glial cell and bulk brain (green) or neuron (blue) **b.** List of all overlapping DMPs between glial cell and bulk brain (green) or neuron (blue). Comparison was performed on CpGs identified in DMR and DMP analyses in bulk tissue and neurons (BS-DMP, cohort 2), respectively, from published studies (Huynh et al. 2014 and Kular et al. 2019).

#### Supplementary Figure 3

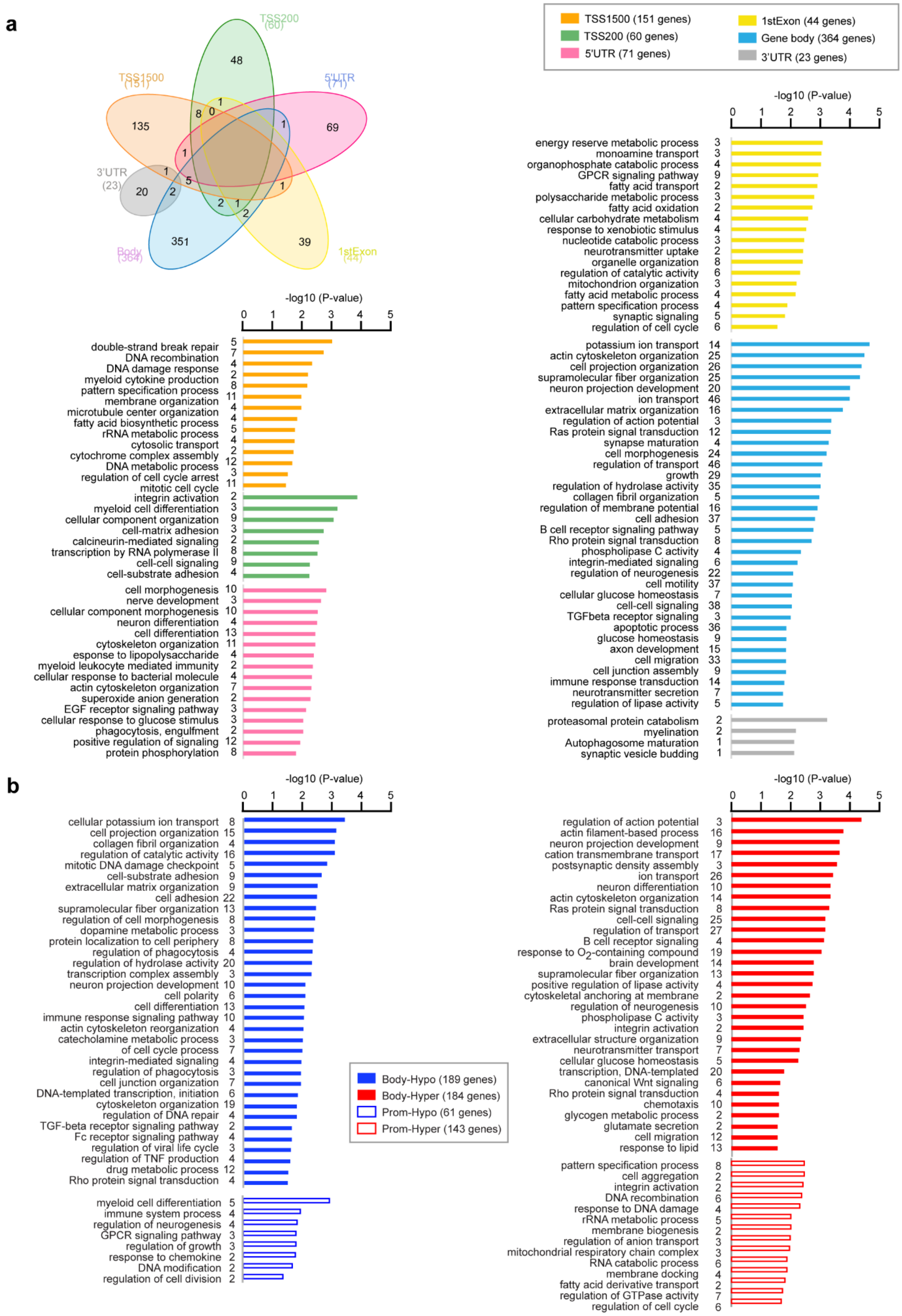

**Supplementary Figure 3.** GO analysis of DMPs according to their genomic location.

**a.** Venn diagram illustrating the overlap between DMPs in different genomic segments, with barplots representing enrichment of GO terms (using overrepresentation analysis) in each genomic feature. **b.** GO analysis of hypomethylated (blue) and hypermethylated (red) DMPs located in gene bodies (filled bars) or in promoters (TSS1500+TSS200, unfilled bars). Enrichment is represented with -log10 (P-value) using overrepresentation analysis. Of note, since fewer DMP-genes were analyzed in some genomic features, results should be taken with caution and the number depicted next to GO terms indicate the number of genes included in each enriched category.

#### Supplementary Figure 4

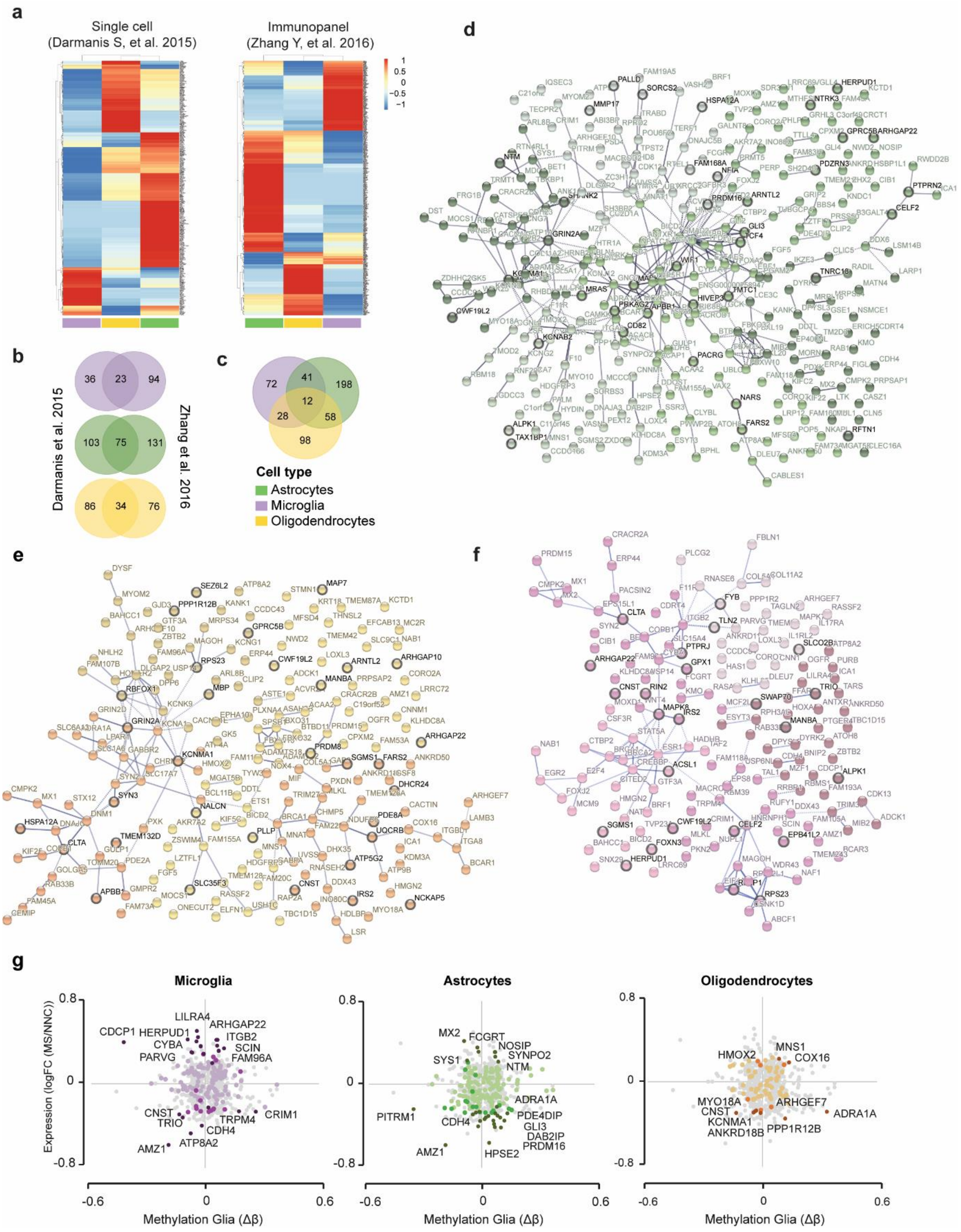


**Supplementary Figure 4.** Annotation of the DMP-genes according to their cellular origin.

**a.** Heatmap illustrating the hierarchical clustering of DMP-gene (P ad < 0.05 in publically available dataset from normal human post-mortem brain transcriptomic analyses (Darmanis *et al.*, 2015; Zhang *et al.*, 2016). Color gradient depicts z score-transformed expression values. **b.** Venn diagram illustrating the overlap of cell type-annotated genes between the two datasets. **c.** Venn diagram illustrating the overlap between astrocyte-, microglia- and oligodendrocyte-annotated (union of genes from the two dataset-specific analyses). **d-f.** Annotation of DMPs genes ($P_{adj}$ < 0.05) according to cell type-specific expression in astrocytes (**d**), oligodendrocytes (**e**) and microglia (**f**) in the healthy brain (Darmanis *et al*., 2015, Zhang *et al*., 2016) using STRING network analysis. Genes reported dysregulated in single-cell transcriptomics from MS vs. NNC brain are highlighted (circled node, bold font). Grey gradient indicated the strength of data support (darker grey representing stronger evidence, dotted line showing lower level of evidence). Colors gradient represent different clusters (kmeans clustering set at 5 clusters). **g**. Association of DNA methylation changes expressed by microglial cells, astrocytes and oligodendrocytes with gene expression data (RNA-seq) in bulk NAWM vs. control WM (Huynh *et al*., 2014). In each panel, the gradient of colors indicates the degree of significance of the differentially expressed genes (dark color reflecting $P < 0.05$, moderate-dark color $P < 0.1$, light color non-significant) in a given cell type while grey colors represent the other genes not annotated for this given cell type.

#### Supplementary Figure 5

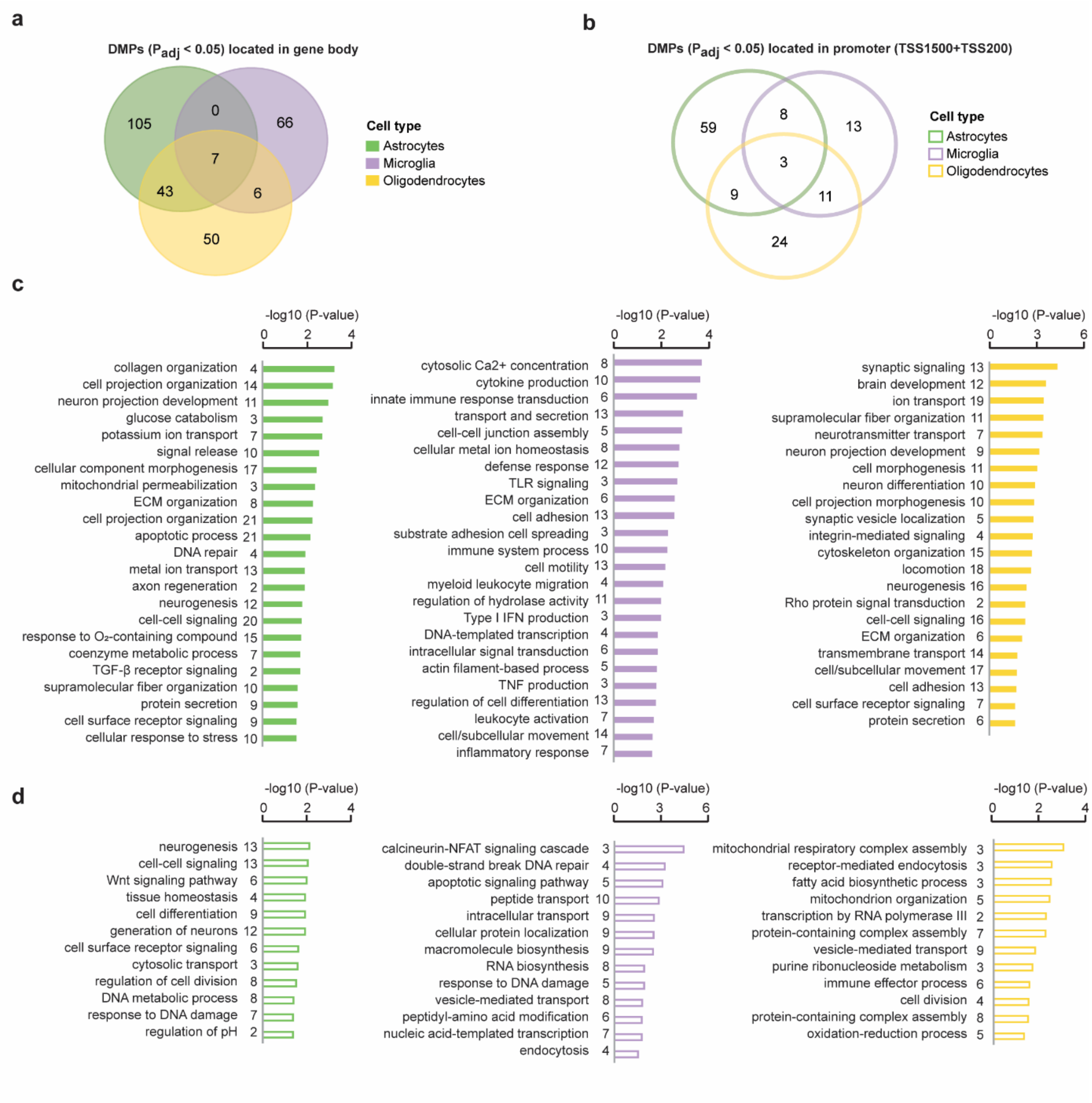


**Supplementary Figure 5.** GO analysis of cell type-annotated DMPs according to their genomic location.

**a-b.** Venn diagram illustrating the overlap between DMPs located in gene body (**a**) and promoter (**b**) annotated as astrocytic (green), microglial (purple) and oligodendrocytic (orange) genes. **c-d.** Enrichment of GO terms from DMPs located in gene body (**c**) or promoter (**d**). Enrichment is represented with -log10 (P-value) using overrepresentation analysis. The number depicted next to GO terms indicate the number of genes included in each enriched category.

### Supplementary Figure 6

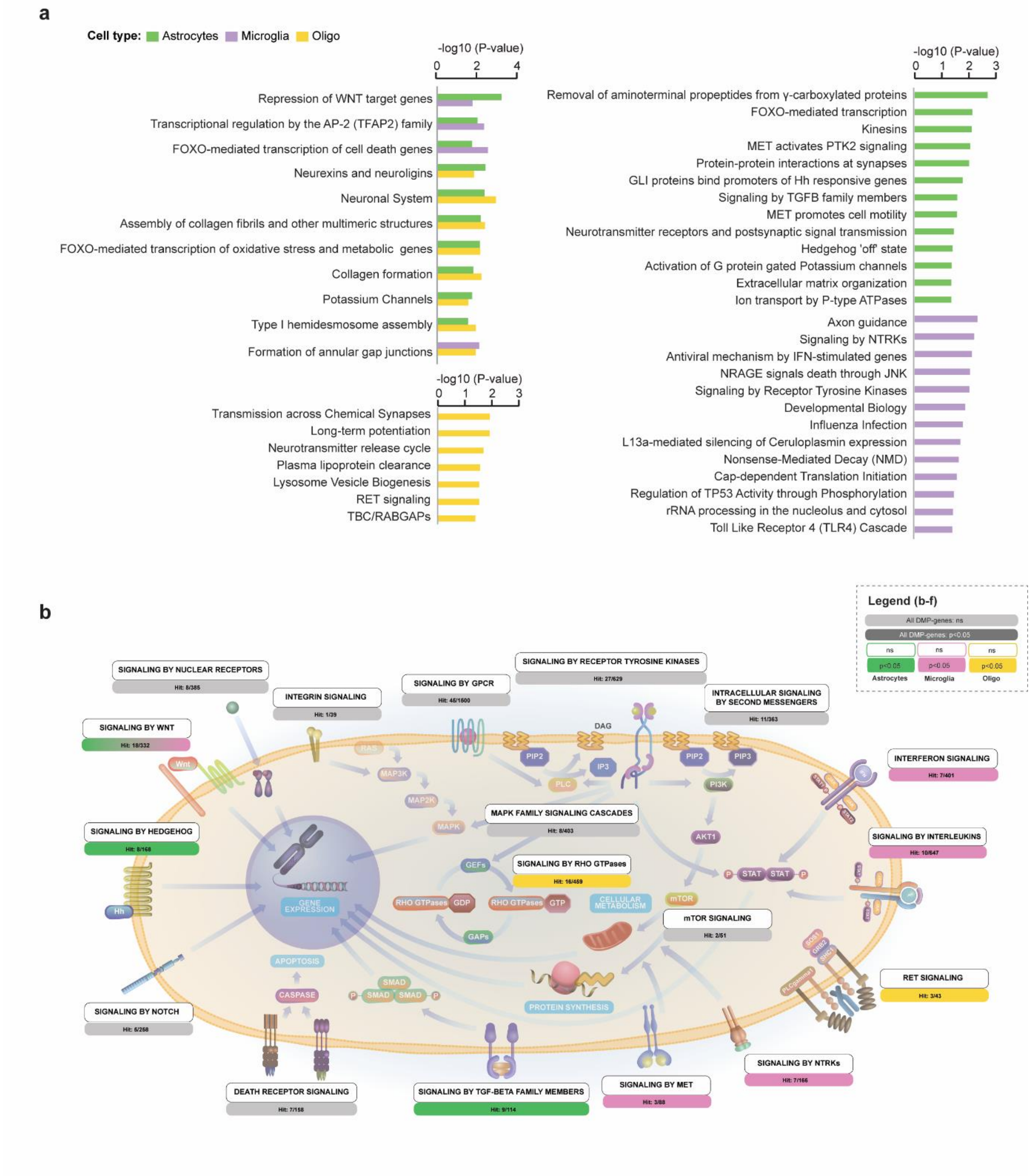


See panel (c-f) in the next page

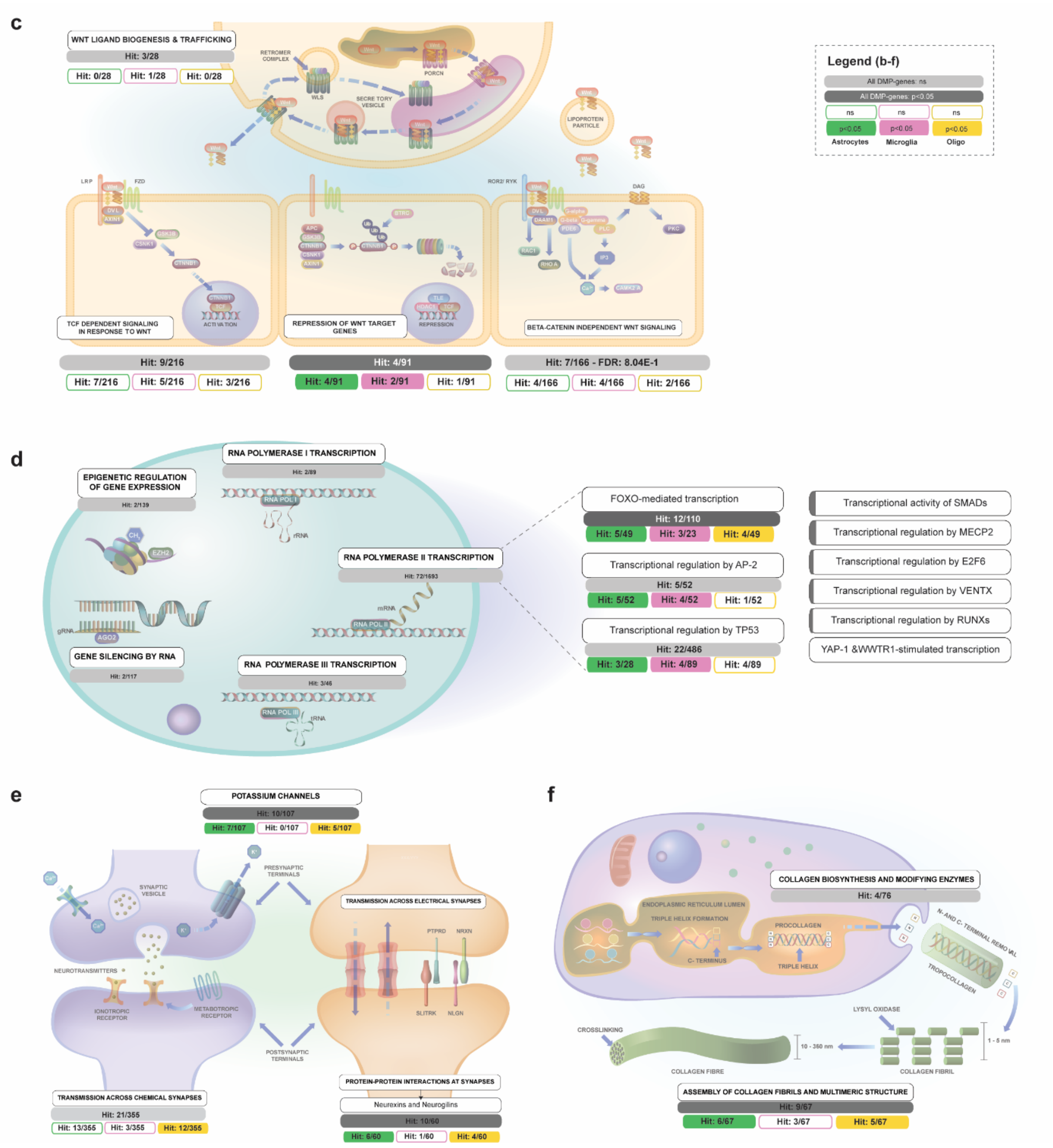


**Supplementary Figure 6.** Pathway (Reactome) analysis of cell type-annotated DMP-genes.

**a.** Shared and unique pathways enriched in glial cells. Enrichment is represented with -log10 (P-value) from Reactome analysis. **b-c.** Schematic illustration of the enriched pathways involved in signal transduction (R-HSA-162582) (**b**), Wnt signaling (R-HSA-195721) (**c**), gene expression (R-HSA-74160) (**d**), neuronal system (R-HSA-112316) (**e**) and ECM organization (R-HSA-1474290) (**f**).

#### Supplementary References


Huynh, J.L. *et al.* Epigenome-wide differences in pathology-free regions of multiple sclerosis-affected brains. *Nat Neurosci* **17**, 121-30 (2014).

Kular, L. *et al.* Neuronal methylome reveals CREB-associated neuro-axonal impairment in multiple sclerosis. *Clin Epigenetics* **11**, 86 (2019).

Darmanis, S. *et al.* A survey of human brain transcriptome diversity at the single cell level. *Proc Natl Acad Sci U S A* **112**, 7285-90 (2015).

Zhang, Y. *et al.* Purification and Characterization of Progenitor and Mature Human Astrocytes Reveals Transcriptional and Functional Differences with Mouse. *Neuron* **89**, 37-53 (2016).

Schirmer, L. *et al.* Neuronal vulnerability and multilineage diversity in multiple sclerosis. *Nature* **573**, 75-82 (2019).

Jakel, S. *et al.* Altered human oligodendrocyte heterogeneity in multiple sclerosis. *Nature* **566**, 543-547 (2019).
